# A generalizable functional connectivity signature characterizes brain dysfunction and links to rTMS treatment response in cocaine use disorder

**DOI:** 10.1101/2023.04.21.23288948

**Authors:** Kanhao Zhao, Gregory A. Fonzo, Hua Xie, Desmond J. Oathes, Corey J. Keller, Nancy Carlisle, Amit Etkin, Eduardo A Garza-Villarreal, Yu Zhang

**Affiliations:** Department of Bioengineering, Lehigh University, Bethlehem, PA, USA; Center for Psychedelic Research and Therapy, Department of Psychiatry and Behavioral Sciences, Dell Medical School, The University of Texas at Austin, TX, USA; Center for Neuroscience Research, Children’s National Hospital, Washington, DC, USA; George Washington University School of Medicine, Washington, DC, USA; Center for Neuromodulation in Depression and Stress, Department of Psychiatry, University of Pennsylvania Perelman School of Medicine, PA, USA; Wu Tsai Neuroscience Institute, Stanford University, Stanford, CA, USA; Department of Psychiatry and Behavioral Sciences, Stanford University, Stanford, CA, USA; Department of Psychology, Lehigh University, Bethlehem, PA, USA; Alto Neuroscience, Inc., Los Altos, CA, USA; Instituto de Neurobiología, Universidad Nacional Autónoma de México campus Juriquilla, Querétaro, Mexico; Department of Electrical and Computer Engineering, Lehigh University, Bethlehem, PA, USA

## Abstract

Cocaine use disorder (CUD) is a prevalent substance abuse disorder, and repetitive transcranial magnetic stimulation (rTMS) has shown potential in reducing cocaine cravings. However, a robust and replicable biomarker for CUD phenotyping is lacking, and the association between CUD brain phenotypes and treatment response remains unclear. Our study successfully established a cross-validated functional connectivity signature for accurate CUD phenotyping, using resting-state functional magnetic resonance imaging from a discovery cohort, and demonstrated its generalizability in an independent replication cohort. We identified phenotyping FCs involving increased connectivity between the visual network and dorsal attention network, and between the frontoparietal control network and ventral attention network, as well as decreased connectivity between the default mode network and limbic network in CUD patients compared to healthy controls. These abnormal connections correlated significantly with other drug use history and cognitive dysfunctions, e.g., non-planning impulsivity. We further confirmed the prognostic potential of the identified discriminative FCs for rTMS treatment response in CUD patients and found that the treatment-predictive FCs mainly involved the frontoparietal control and default mode networks. Our findings provide new insights into the neurobiological mechanisms of CUD and the association between CUD phenotypes and rTMS treatment response, offering promising targets for future therapeutic development.

## INTRODUCTION

Cocaine use disorder (CUD), a disorder marked by compulsive cocaine use, affects over one million people in the United States according to the 2019 national survey on drug use ^1, 2^. Despite the availability of clinical psychosocial treatments for CUD, the high relapse rate highlights the need for more effective treatments for all CUD patients ^3^. A robust and generalizable phenotyping neuroimaging-based biomarker could be helpful to advance our understanding of the neurobiological mechanism and pathophysiology of CUD ^4, 5^. Moreover, compared to diagnostic questionnaires, such phenotyping biomarkers could be critical in forensic settings by providing an objective means to reduce controversy in evaluations of mental insanity and minimize errors in detecting malingering ^6^.

Magnetic resonance imaging (MRI) has been a useful tool for detecting the abnormalities in brain structure and activity associated with CUD. Structural MRI studies using voxel-based morphometry have found reduced gray matter volume in brain regions such as the orbitofrontal cortex, anterior cingulate cortex, and superior temporal cortex in CUD patients compared to healthy controls ^7^. On the other hand, by examining brain activity from blood-oxygen-level-dependent (BOLD) signals during cocaine craving, functional MRI (fMRI) studies have observed stronger activation in the left dorsolateral prefrontal and bilateral occipital cortex of CUD patients than healthy controls ^8^.

Compared to the analyses of brain structural alternations or regional activations, functional connectivity (FC) measures the neural coupling across various brain regions and can be a more sensitive indicator of brain dysfunction than morphological features and task-related brain regional activation ^10, 11^. For instance, dysfunctions in neural circuits involving thalamus were observed in episodic migraine patients despite normal anatomical structure ^12^ and FC involving the prefrontal cortex changed in anticipation of future spatial and letter tasks rather than prefrontal cortical activation in adults with focal brain lesions^13^.

Thus, increasing studies have focused on examining functional connectivity abnormalities in CUD. It has been found that the resting-state functional connections involving the prefrontal cortex of CUD patients are significantly weaker than healthy controls ^9^.Additionally, resting-state FCs involving the frontoparietal control network (FPC) and default mode network (DMN), have been found to contribute largely to the classification of CUD patients versus healthy controls ^14^. Despite progress made by existing fMRI-based CUD phenotyping studies, their utility remains limited due to small sample sizes (no more than 80 subjects) ^9, 14–17^ and a lack of independent validation ^14, 18, 19^, resulting in likely overestimated accuracy.

Recent studies have found associations between phenotyping biomarkers and clinical outcomes in some typical mental disorders ^20^. For example, elevated presynaptic striatal dopaminergic function, a phenotype of schizophrenia, was shown to predict response to treatment of psychotic symptoms ^20, 21^.

Abnormally increased FC within DMN, a depression risk indicator, has been observed to decrease with repetitive transcranial magnetic stimulation (rTMS) treatment ^22, 23^. In the case of CUD patients, albeit a lack of effective therapy, abundant studies have observed a significant reduction in cocaine craving following repetitive transcranial magnetic stimulation (rTMS) ^24, 25^. Uncovering the association between CUD phenotyping FCs and rTMS treatment response can potentially advance our understanding of neurobiological mechanisms underlying the disease. Furthermore, phenotyping biomarkers linked to treatment outcomes could be used as targeted regions in brain pathways for novel therapeutic intervention. However, such critical association remains underexplored, partially due to limited exploration of reliable phenotyping biomarkers.

In this study, we sought to address the aforementioned limitations by establishing a robust and replicable FC signature to accurately characterize neural circuit abnormalities in CUD phenotyping and reveal its association with rTMS treatment response. We first aimed to train a classification model to obtain a generalizable FC phenotype for distinguishing CUD patients from healthy controls using the XGBoost algorithm ^26^, a robust ensemble learning technique known for mitigating overfitting.

Specifically, we utilized FC features extracted from resting-state fMRI of a discovery cohort comprising 71 CUD patients and 58 healthy controls to build a machine learning model, using XGBoost combined with data augmentation and feature selection, to identify a discriminative FC signature. We demonstrated the robustness of the model by 10-fold cross-validation in the discovery cohort and further confirmed its generalizability through an external validation using an independent cohort consisting of 81 CUD patients and 82 healthy controls. Additionally, we conducted statistical analyses to investigate the associations between the identified discriminative FCs and substance use history as well as clinical assessments (eg. non-planning score in *Barratt Impulsiveness Scale*). Furthermore, we aimed to investigate the link between the FC signature and treatment outcome, which was typically missed in previous phenotyping studies ^20, 27, 28^. By examining the discriminative FC signature as a predictor of treatment response to rTMS in 45 CUD patients who were randomly assigned to receive either 5 Hz active or sham rTMS on the left dorsolateral prefrontal cortex, we demonstrated the utility of this diagnostic FC signature for prognostic purpose of predicting treatment response.

## RESULTS

### Functional Connectivity Signature Distinguishes CUD from Healthy Controls

Regional FC features were calculated from resting-state fMRI of 71 CUD patients and 58 healthy controls, using Schaefer parcellation with 100 parcels^29^. Then we built up the XGBboost classification model, utilizing the abnormal FCs which significantly differentiated CUD patients and healthy controls (two-simple t-test p < 0.05). The FC-based XGBoost model showed promising results for the cross-validated phenotyping of CUD with an accuracy of 0.83 ± 0.10, sensitivity of 0.80 ± 0.18, and specificity of 0.85 ± 0.10. (Figure 1A). To understand the contribution of each FC feature to the classification, we calculated the frequency of the feature appearing in all trees of the XGBoost model and plotted the most discriminative FCs for better visualization (Figure 1B). All discriminative FCs were shown in Figure S1. The most discriminative FCs were between the cuneus and precuneus and between the middle cingulate cortex and superior temporal cortex. We further investigated the network-level connectivity that was critical for the CUD classification by averaging the FC discriminative pattern according to Yeo’s 7 networks ^30^ (Figure 1C). Prominent discriminative FCs were observed between DMN and limbic network (LIM), between FPC and somatomotor network (SMN), between visual network (VIS) and dorsal attention network (DAN), and between FPC and ventral attention network (VAN). The contribution of these four network-level connections to the model performance was confirmed by 1000 random permutations for each pairwise network connection (Figure S3B). The network-level feature importance, defined as the average feature importance within a network and between the network and all other networks, further indicated that CUD classification was mainly driven by between-network connections involving the FPC, DMN, DAN, and LIM (Figure 1D).

**Figure 1.**
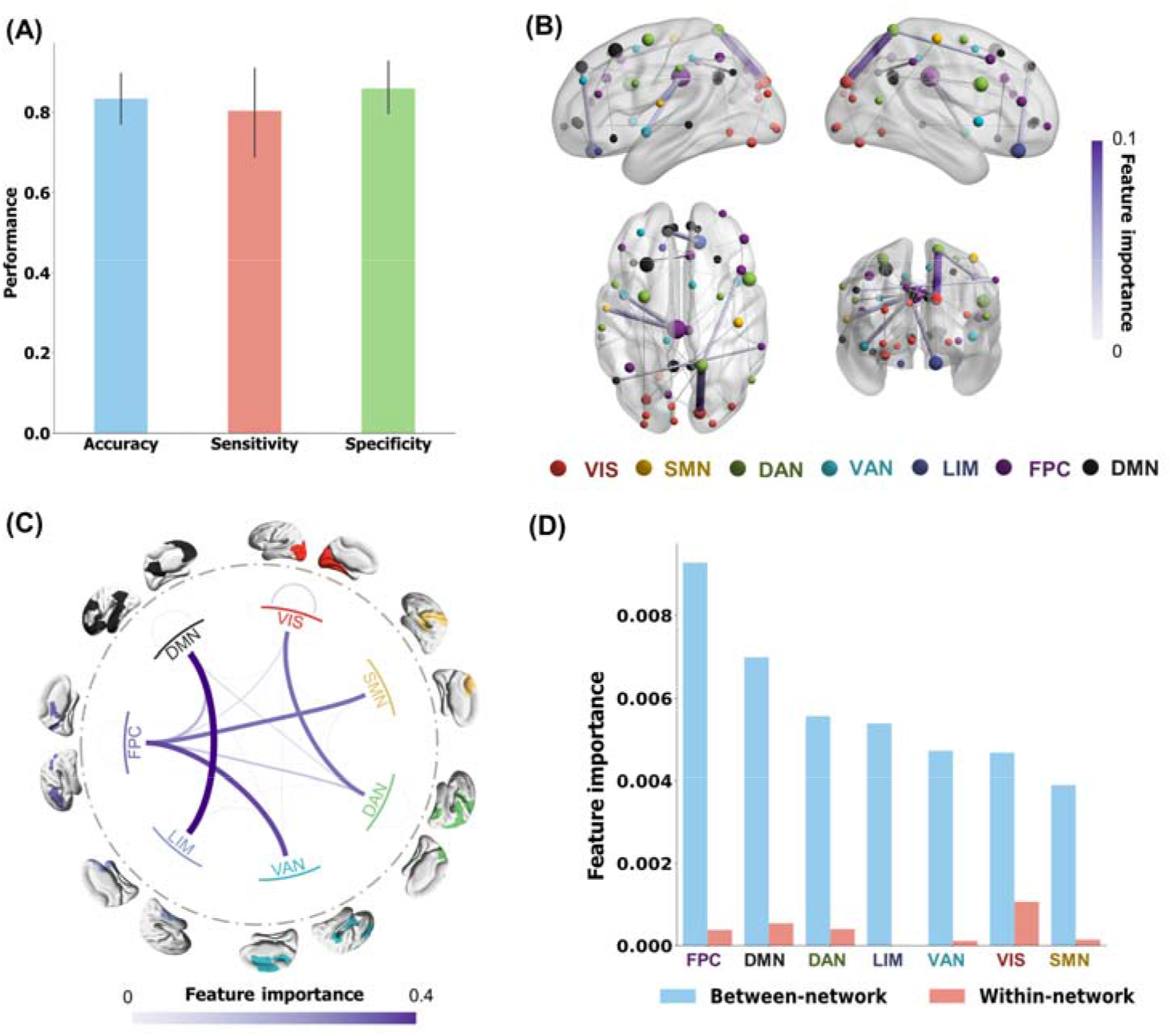
FC phenotype of CUD patients versus healthy controls. **(A)** Classification performance across 10-fold cross-validations: the accuracy, sensitivity, and specificity of the FC-based XGBoost model are 0.83 ± 0.10, 0.80 ± 0.18, and 0.85 ± 0.10, respectively. **(B)** Visualization of the 40 most discriminative FC features as identified by the XGBoost model, by calculating frequency of the features appearing in all the trees of the model. Node size indicated the node strength calculated from the sum of the linked FC importance. **(C)** Network-level discriminative pattern obtained by grouping FC importance based on Yeo’s 7 networks. **(D)** Averaged between-network and within-network FC strengths. The between-network FC strength was calculated by averaging the importance of discriminative connections across each network and all other networks. VIS, visual network; SMN, somatomotor network; DAN, dorsal attention network; VAN, ventral attention network; LIM, limbic network; FPC, frontoparietal control network; DMN, default mode network.

To explore the directionality of the discriminative FCs, we conducted two-sample t-tests on them between CUD patients and healthy controls, as shown in Figure 2. These FCs were divided into hyperconnections, where FCs were significantly higher in the CUD group compared to the healthy controls group, and hypoconnections, where FCs were significantly lower in the CUD group. For better visualization, the top 40 hyperconnections and hypoconnections were shown. The most important hyperconnections were mainly located in the middle cingulate cortex, precuneus and inferior frontal cortex (Figure 2A, Figure S2A). Some of the most significant hyperconnections included connections between FPC and VAN as well as between VIS and DAN. The most important hypoconnections were primarily found in the posterior cingulate cortex, superior medial frontal cortex and orbital frontal cortex (Figure 2B, Figure S2B). The most prominent hypoconnections were found between DMN and LIM.

**Figure 2.**
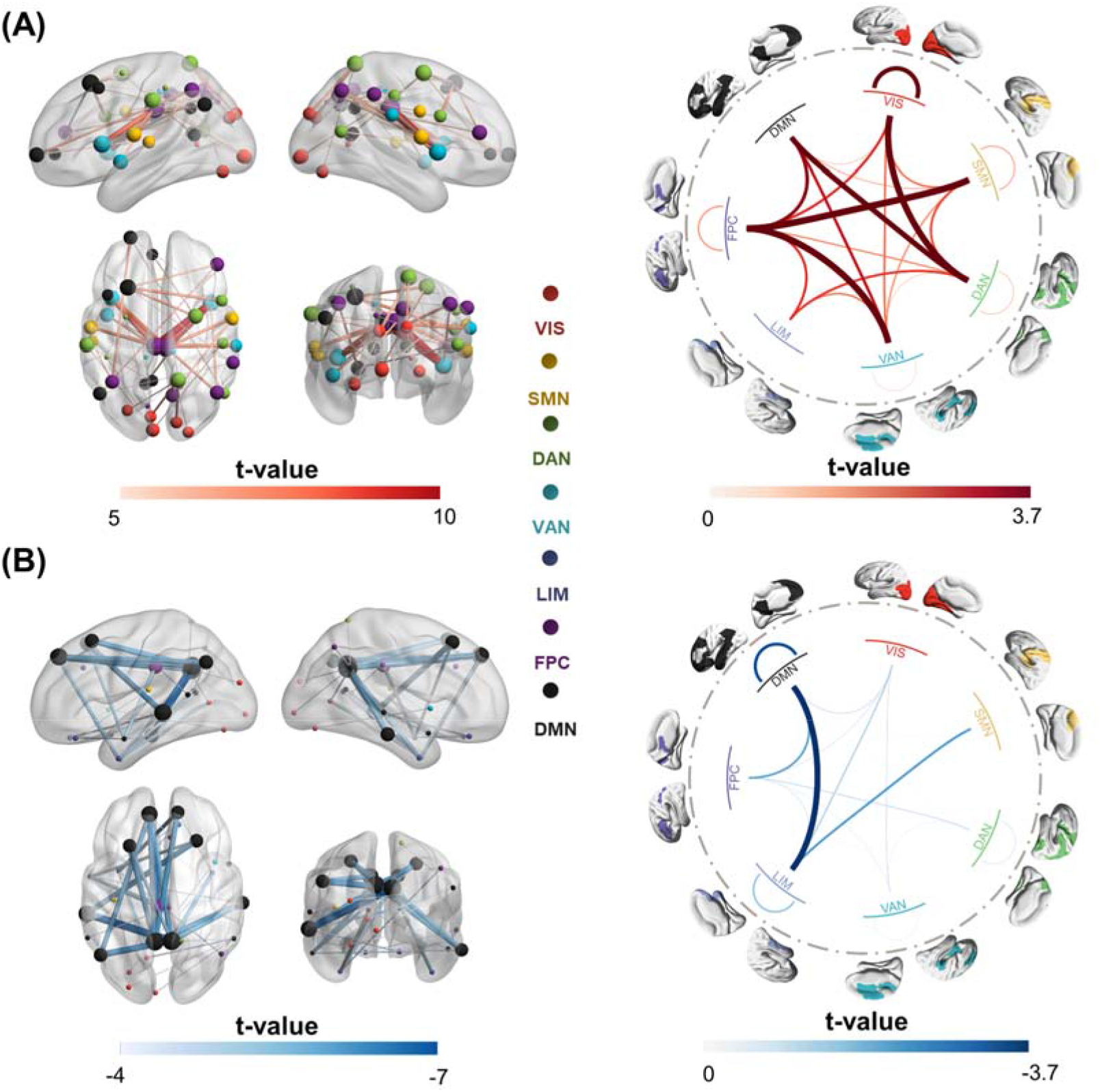
Statistical difference in the classifier-identified discriminative FCs between CUD patients and healthy controls, examined by two-sample t-tests. **(A)** The 40 most significant hyperconnections (CUD > healthy controls), with their t-values shown in the left panel. Node size indicated the node strength calculated from the sum of the linked FC t-value. The right panel shows the average t-values across all significant hyperconnections, calculated at the network level. **(B)** The 40 most significant hypoconnections (CUD < healthy controls), with their t- values shown in the left panel. Node size indicated the node strength calculated from the sum of the linked FC t- value. The right panel shows the average t-values across all significant hypoconnections, calculated at the network level. Only the FCs that passed the FDR correction (p<0.05) are considered significantly discriminative FCs.

### Association Between Discriminative Connections and Clinical Assessments

To investigate the relationship between CUD-predictive connections and clinical assessments, we first applied two-sample t-tests to compare the discriminative network-level connections between subjects with other disorder diagnostic labels versus healthy controls. The statistical analyses were focused on the top 4 discriminative network connections since they were significantly more important for the classification than other network connections (Figure S3A, an inflection point was observed in the 4th network connection; Figure S3B, these four connections significantly contribute to the classification performance evaluated by 1000 permutations). The abnormal connections between DMN and LIM and between VIS and DAN were found to be significantly related to drug use histories, including hallucinogens, stimulants, and inhalants (Table S1). When other substance use disorder and drug use history variables recorded in MINI were regressed out, the model still exhibited a decent performance (Figure S4, accuracy = 0.71 ± 0.14, sensitivity = 0.67 ± 0.20, and specificity = 0.74 ± 0.14). The discriminative signature was generally consistent (Figures S4 and S1, the Spearman correlation between these two signatures was r = 0.38, p = 6.12 × 10^-171^ and the top 10 discriminative FCs in both signatures were highly overlapping). These findings indicated that the discriminative FC was mainly driven by cocaine addiction, and also confounded with polysubstance use history. We then examined the correlation between the discriminative network-level connections and other continuous clinical assessments (Pearson’s correlation for normally distributed continuous variables, e.g. subscales of the Barratt Impulsiveness Scale (BIS), and Spearman’s correlation for non-normally distributed continuous variables, e.g. subscales of the World Health Organization Disability Assessment Schedule (WHODAS)) (Table S2). The p-values of each connection across all those clinical variables were corrected for FDR. The connections between VIS and DAN were positively correlated with non-planning impulsiveness. The connections between VAN and FPC and between SMN and FPC showed positive correlations with various psychological symptoms such as depression and anxiety in Symptom Checklist-90-revised. Furthermore, the connections between DMN and LIM, between DAN and VIS were significantly correlated with the consuming pattern of tobacco (Table S3).

### Functional Connectivity Signature Generalizes to an Independent Cohort

To test the generalizability of our findings, we applied the cross-validated classifier trained with the discovery cohort to replication cohort including 81 healthy controls from the UCLA-CNP dataset and 82 CUD patients from both the NYU and SUDMEX-TMS datasets (See more details about these datasets in Online Methods section). The results show that our classification model still achieved decent performance (Figure 3A, accuracy = 0.70, sensitivity = 0.69, and specificity = 0.70). We used Combat^31^ for site correction based on the demographic information to alleviate site effect caused by different MRI scanning parameters on the replication cohort. However, such site effect might still affect the validation performance. To investigate whether and how the validation performance of phenotyping was influenced by the site information, we applied the well-trained CUD classifiers based on the discovery cohort to identify site labels of all subjects in independent cohort from either the UCLA-CNP (81 healthy controls and 145 patients with other mental disorders), or the combination of NYU and SUDMEX-TMS datasets. If the site contributed largely to the replication phenotyping performance, the models should be able to classify site labels with decent performance. The performance of site classification (Figure S5A, accuracy = 0.58; sensitivity = 0.53; specificity = 0.72; p_permute_ = 0.001) was significantly but marginally higher than the chance level. To control the sample size effect from different sites, we down-sampled the UCLA-CNP equal to the size of NYU and SUDMEX-TMS 1000 times and reclassified the site labels. The mean performance in (Figure S5B, accuracy_mean_ = 0.61; sensitivity_mean_ = 0.53; specificity_mean_ = 0.70) was similar to the site classification using all subjects in UCLA-CNP (Figure S5A). Importantly, the phenotyping classification accuracy was significantly higher than the subsampling site classification accuracy (p < 0.001), which suggested the reproduced phenotyping performance can be derived mostly from the diagnostic label instead of site labels. To further demonstrate that the replicated phenotyping performance was not primarily driven by site effect, we employed an alternative strategy. We trained a classification model using the independent replication cohort and then tested its predictive ability on the discovery cohort. If the performance of classification was dominantly attributed to site effect, the model should not be able to accurately classify CUD from healthy controls in the discovery cohort. As shown in Figure S6, the ten-fold cross-validation on the independent cohort yielded an accuracy of 0.77, sensitivity of 0.78, and specificity of 0.75. When applying the trained classifier to the discovery cohort, we observed that the accuracy, sensitivity, and specificity in the discovery cohort were 0.63, which was significantly larger than permutation test (p < 0.001). This again demonstrated that the phenotyping performance was not dominantly derived from the site effect.

**Figure 3.**
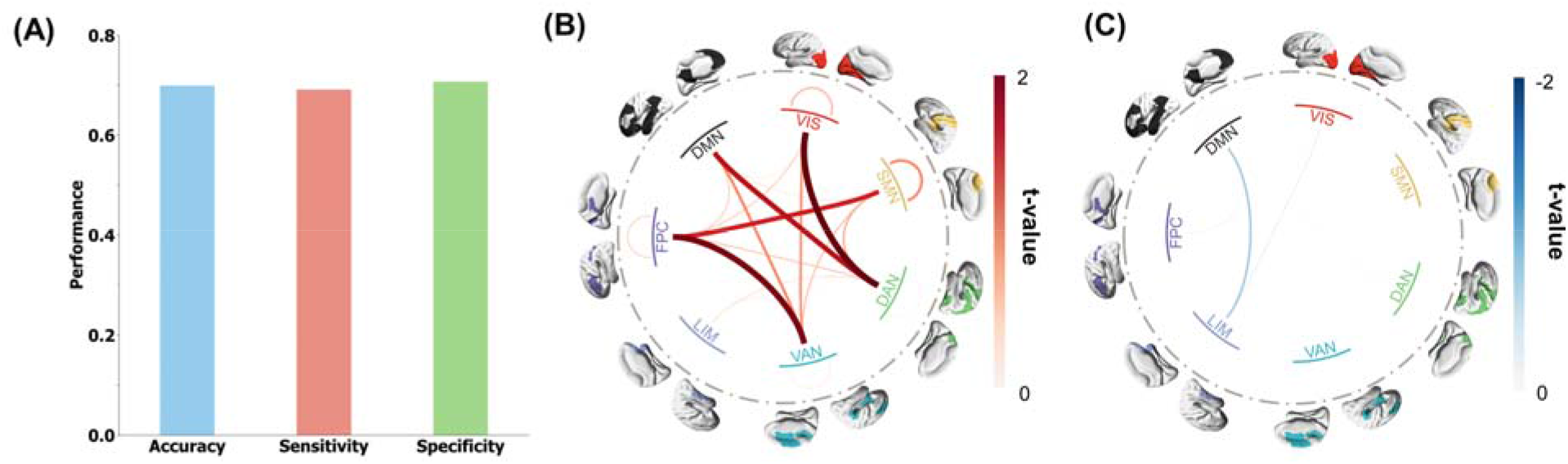
Replication of the discriminative FC signature in the independent cohort. **(A)** The accuracy, sensitivity, and specificity of predicted results in the independent replication cohort, using predictive FC signature obtained from the discovery cohort. **(B, C)** Replication results of the two-sample t-test analysis that compares the network-level predictive FCs in CUD patients and healthy controls showing the network-level hyperconnectivity (**B**) and network-level hypoconnectivity (**C**).

Next, we sought to verify that the predictive biomarker generalized to the independent replication cohort is due to FC abnormalities consistent with those identified in the discovery cohort. Two-sample t-tests were conducted to detect differences in the predictive FCs between CUD patients and healthy controls. All hyper-connections and hyper-connections were averaged into seven networks (Figure 3B). The network hyper-connections were dominated by connections between FPC and VAN; between VIS and DAN. The network hypo-connections were dominated by connections between DMN and LIM (Figure 3C). A similar difference in predictive FCs between CUD patients and healthy controls in the discovery cohort and validation cohort was observed (Pearson correlation = 0.64, p = 2.19 × 10^-84^). Such consistent FC difference observed between the discovery cohort and validation cohort might explain the highly reproducible performance.

### Predicting TMS Treatment Response Using Discriminative FCs

Next, we sought to test whether the discriminative FC signature could also serve as a prognostic biomarker to predict treatment response. To this end, we utilized the FCs calculated from baseline resting-state fMRI of CUD patients who were randomly assigned rTMS treatment in the clinical trial cohort (SUDMEX-TMS dataset, 20 sham rTMS and 25 acitve rTMS). In the active rTMS treatment phase, the stimulation with 5000 pulses at 5 Hz per-day was delivered to the left dorsolateral prefrontal cortex. For the patients in active rTMS treatment arm, a relevance vector machine model ^32^ was then trained with these selected FCs to predict changes in the visual analog scale (VAS) score which is designed to evaluate the participants’ craving of cocaine ^33^. To determine the relationship between the discriminative FC features and treatment response to rTMS, we used the identified connectivity signature as a mask to select the FCs of baseline CUD patients who were assigned rTMS treatment in the clinical trial cohort (SUDMEX-TMS dataset). The prediction performance was evaluated by Pearson’s correlation and r-squared value (R^2^) between the predicted VAS score changes and the observed ones using ten 5-fold cross-validation. The results show that the CUD discriminative FC features have decent predictive ability (r = 0.59, p = 0.002, p_permute_ = 0.002) for the response to active rTMS treatment (Figure 4A). Furthermore, applying these models trained on patients assigned to the active rTMS arm to those assigned to the sham rTMS arm failed to predict treatment outcome (r = 0.36, p = 0.10, p_permute_ = 0.07, Figure 4B), indicating the discriminative features were specifically predictive for response to the active rTMS treatment. The statistical comparison of the model performance of ten 5-fold cross-validated models demonstrated the significance of specificity (Wilcoxon signed-rank test of prediction performance of ten 5-fold models in two treatment arms, w_R_2 = 3.78, p_R_2 = 0.0002; w_r_ = 3.40, p_r_ = 0.0007, Figure 4C). To further confirm the significance of relationship between our discriminative FCs and the response to active rTMS treatment, we repeatedly evaluated the prediction performance 1000 times after randomly subsampling all FCs with equal size to discriminative FCs. Meanwhile, we evaluated the prediction performance in ten 5-fold cross validation, training with all FC features. The result shows that the identified discriminative FCs outperformed all FCs (Figure S7A, the performance of the ensemble ten models, r = 0.42, p = 0.037, R^2^ = 0.07; Figure S7B, Wilcoxon signed-rank test of the performance difference between training using phenotyping FC and all FC. w_R_2 = 3.63, p_R_2 = 0.0003; w_r_ = 3.55, p_r_ = 0.0004.) and the randomly subsampled FCs in predicting active rTMS treatment (Figure S7C, the p value of the performance difference between training with phenotyping FC and random-subsampling FC was 0.02). These results demonstrated that the rTMS treatment response information was enriched in phenotyping FCs, compared to random-subsampling FCs and whole FCs.

**Figure 4.**
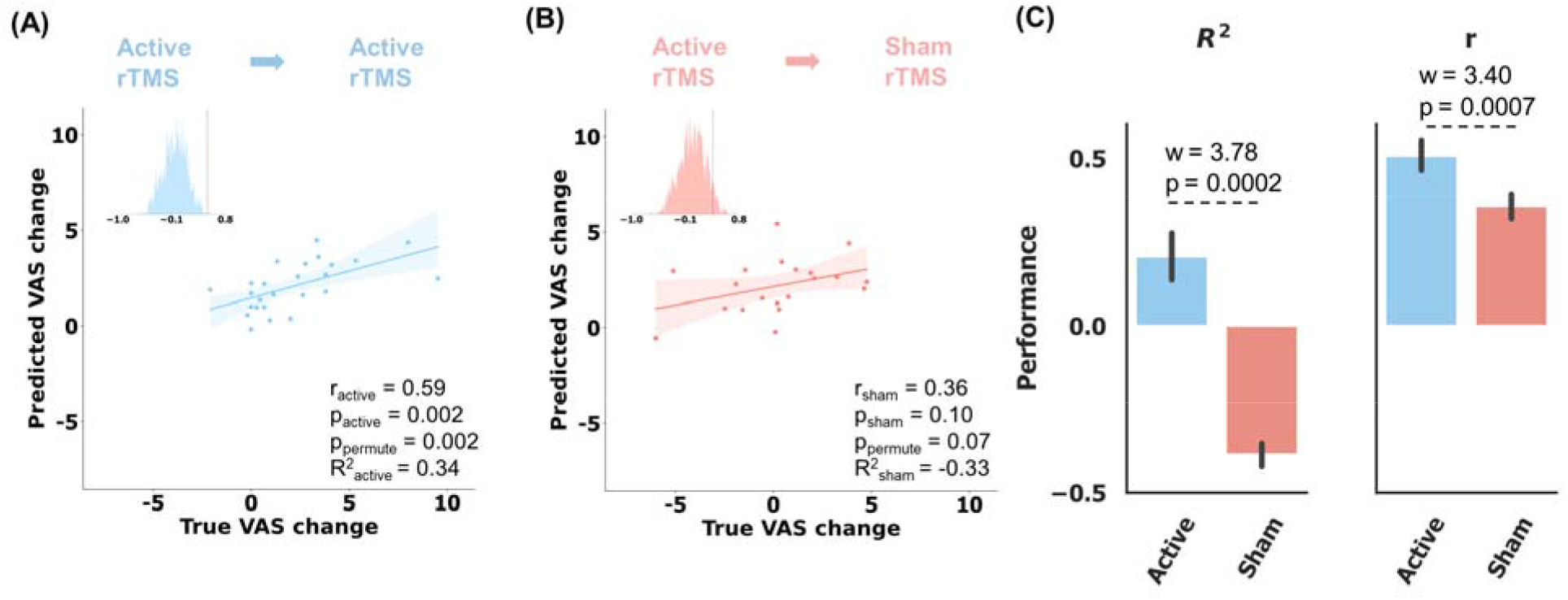
Prediction of the craving visual analog scale (VAS) score changes specific to active rTMS in ten 5- fold cross-validation. The statistical significance of prediction performance was confirmed by 1000 random permutation tests. **(A)** Scatter plot showing the prediction results of all patients who were assigned active treatment, using the CUD discriminative FC features (r = 0.59, p = 0.002, ppermute = 0.002). The predicted scores were averaged from the scores predicted from ten 5-fold models. **(B)** Application of active rTMS-trained models to the sham rTMS failed to predict the clinical score change (r = 0.36, p = 0.10, ppermute = 0.07). **(C)** Difference in the prediction performance between active and sham rTMS treatment groups. It was evaluated by Wilcoxon signed-rank test of all prediction performance of ten 5-fold models. w_R^2^_ = 3.78, p_R^2^_ = 0.0002; w_r_ = 3.40, p_r_ = 0.0007.

We then visualized the importance of the CUD-discriminative FCs in predicting active rTMS treatment response in Figure S8A and further averaged those FCs’ importance into Yeo’s 7 networks in Figure S8B. The predictive FCs dominantly came from the frontal lobe involving DMN, LIM, and FPC. However, the treatment response predictive connectivity pattern was not similar to the phenotyping pattern (Figures 1C and S1). As shown in the Venn diagram (Figure S9G), all discriminative and abnormal FCs (The hyperconnections and hypoconnections in Figure S2) contributed to the treatment outcome prediction and 74 of the top 100 treatment predictive FCs were captured in CUD discriminative and abnormal connectivity signature. The correlation between the top rTMS outcome predictive FCs and the VAS scores change and these two FC distributions across two cohorts as a visualization example indicated that the top rTMS predictive FCs were with moderate abnormality and CUD discriminative importance (Figure S9A-F).

## DISCUSSION

In this study, we have developed a robust FC-based classification model for CUD phenotyping using resting-state fMRI. Our analyses demonstrated its suitable accuracy, sensitivity, and specificity in classifying CUD patients and healthy controls. The high classification performance was further validated in an independent replication cohort, making this study the first to identify a generalizable FC signature to distinguish CUD from healthy controls based on neural circuit abnormalities. The statistical analysis of difference in distinguishable FCs between CUD patients and healthy controls provided evidence of the network-level connectome abnormalities in CUD, primarily involving the FPC, DMN, DAN, and VAN. Additionally, the association between our identified discriminative FCs and various clinical assessments showed that the abnormal FCs of CUD were correlated with substance use history and psychiatric symptoms such as depression and impulsivity. Importantly, the CUD-discriminative FCs demonstrated the capacity to predict treatment response to rTMS. These results together suggest that our identified FC signature could serve as a novel diagnostic tool to assist with CUD diagnosis, offer new insights into the neuropathology of CUD, and potentially predict course of treatment with rTMS.

### Discriminative Hyperconnectivity and Hypoconnectivity

Previous studies of the brain connectome in individuals with CUD have consistently shown hyperconnections in key regions involved in response inhibition, such as the middle cingulate cortex, anterior cingulate cortex of FPC ^34^, inferior frontal cortex of DAN ^35^ and insular cortex of VAN ^35, 36^.

These regions form the canonical response inhibition circuit in the brain^37^. It is aligned with the hypothesis that chronic cocaine use is associated with a decline in inhibitory control function of the cortex^38^. In particular, the inferior frontal cortex is known for its role in inhibitory control ^39^. Studies have reported low fractional anisotropy in white matter and elevated Go/NoGo response-inhibition task-related brain activation of the inferior frontal cortex in CUD ^40, 41^. Consistent with these previous findings, our study identified a positive correlation between the connections involving DAN, VIS, and impulsiveness (loss inhibition), especially non-planning impulsiveness (Table S2). Moreover, the connectivity between FPC and VAN was found to positively correlate with various neuropsychological symptoms. This is in line with the observation that cognitive dysfunction and emotional dysregulation are mediated by FPC and VAN in disorders such as depression, anxiety, and autism ^42^.

Furthermore, our findings suggested that the reduced connectivity between DMN and LIM played a critical role in the classification model for phenotyping CUD. The key regions involved in the between-network connectivity were the posterior cingulate cortex, superior medial prefrontal cortex, orbital frontal cortex, and temporal pole. Previous research has shown that the temporal pole–medial prefrontal cortex circuit ^43^ and activation of posterior cingulate cortex following various cocaine cue stimuli ^44^ were highly predictive of cocaine relapse. These regions in DMN and LIM play a key role in social emotion perception and regulation of negative emotion ^43, 45–47^. This was in line with our results that the FC between DMN and LIM (prior to FDR correction) negatively correlated to the severity of individuals’ cognitive impairments in communication and thinking activities assessed by WHODAS ^48^ (Table S2). In addition, cognitive dysfunction has been found to be associated with anxiety severity ^49, 50^. The association was also observed in our result that the reduced FCs between DMN and LIM were negatively correlated with anxiety symptoms (Table S2). Overall, the association between DMN-LIM hypoconnectivity with both cognitive complaints as well as anxiety suggested that future research test these networks as potential sites of intervention for addressing these symptoms in CUD ^51, 52^.

### Association Between CUD and Other Substance Use Disorders

Our study indicates that the connectivity abnormalities in CUD patients are intertwined with other substance use histories. We observed that the phenotyping signature could robustly differentiate the CUD patients with a history of polysubstance use or pure CUD from healthy controls. The intertwined effect from other substance use confounders enhances the model performance with an increased 13% precision. This improvement may be attributed to the shared dysfunction across the brain networks of the VAN, DMN, FPC of polysubstance use disorders ^53–55^. Given the cognitive, emotional, and attention control functions of DMN, FPC, and VAN, it is reported that excitability, impulsivity, aggression, and attention deficits were observed in fetal growth exposed to tobacco, stimulants, or other drugs during pregnancy ^56^. It is congruent with the association between the most CUD-discriminative FCs and other substance use histories. For example, the connections between DMN and LIM, and between VAN and FPC were found to differ between healthy controls and individuals with histories of suicide tendencies, alcohol abuse, other drug use, and tobacco use (Tables S1 and S3). Additionally, the dysfunction of DMN, FPC and ventral attention networks in CUD patients highlights the role of the triple network model across major psychiatric disorders ^42^.

### Association Between CUD-discriminative Signature and rTMS Treatment Response

Our study found that the CUD-discriminative connectivity signature was also significantly and specifically associated with the response to active rTMS treatment. Despite the connectivity pattern linked to treatment response was inconsistent to the CUD-discriminative signature, most rTMS predictive FCs are able to differentiate CUD from healthy controls. This finding might align with the recently proposed core abnormal FC (CA-FC) treatment remission hypothesis ^57^, which assumed that a CA-FC set encompassed all possible abnormal FCs of a disorder (In the reference, they focused on major depressive disorder). Different treatments may specifically target different CA-FC subsets. As revealed from our result, the CA-FC of rTMS treatment primarily came from the frontal lobe involving DMN, LIM, and FPC. It could be congruent with the cocaine molecular pathway. Cocaine affects frontostriatal transmission by blocking dopamine transporters and increasing free dopamine in the nucleus accumbens^58^. This in turn seems to produce an increase in GABA in the prefrontal cortex, increasing limbic system activity by local inhibition ^59^. TMS is thought to indirectly regulate these neurotransmitters and functional connectivity in orbitofrontal, dorsolateral prefrontal and ventromedial prefrontal cortex to reduce craving ^60^, mediating psychological processes such as risk-reward decision-making ^61^. In light of these findings, the prediction power derived from the CA-FC involving frontal lobe across three networks underscores the role of frontostriatal circuit in mediating CUD symptoms and its importance in influencing efficacy of circuit-focused interventions for CUD.

### Limitations

The present study has several limitations and potential areas for improvement that are important to consider. One limitation is the imbalance in gender representation, as more males than females were included in the discovery and replication cohorts, which may result in a bias in our identified signature towards males. Although illicit cocaine use is significantly more prevalent among males than females ^62^, it is interesting to explore whether there is some different brain dysfunction among males and females.

Additionally, the site effect might contribute a bit to the high replication performance since the CUD patients and healthy controls of the replication cohort were obtained from different datasets and the number of patients assigned rTMS treatment was limited. This highlights the need for a better- harmonized data acquisition protocol and the recruitment of more independent subjects in future studies to verify the robustness of our connectivity signature findings and the association to rTMS treatment response. Moreover, most of the CUD patients in the discovery cohort had a history of other drug or alcohol abuse. In the future, it would be valuable to study patients addicted to other substances such as alcohol with cocaine use history to further verify the shared brain dysfunction across different substance use disorders. Lastly, it would be interesting to investigate the progression of brain dysfunction in individuals who used cocaine only limited times without addiction compared to those who developed a cocaine addiction.

## Conclusions

In this study, we built a XGBoost based machine learning model, trained with resting-state fMRI FC to accurately distinguish CUD patients with polysubstance use history from healthy controls in a discovery cohort. We identified a robust FC signature and successfully replicated the finding in an independent cohort, making it a generalizable phenotyping biomarker for CUD. In the FC signature, we found hyperconnectivity between VIS and DAN, and between FPC and VAN, as well as hypoconnectivity between DMN and LIM in both cohorts. These brain network connections were linked to other substance use histories and cognitive dysfunction in depression and impulsivity domains. These results emphasized the dysfunctional links of the six typical brain networks in neuropathology of cocaine use disorder comorbid with other substance use histories. More crucially, the association between the CUD-discriminative signature and response to rTMS treatment was further discovered, highlighting the role of prefrontal cortex across LIM, DMN, FPC in the CUD craving remission progress. The FCs across these three networks have the potential to serve as a decision-making tool for clinical psychiatrists.

## Online Methods

### Discovery Cohort

#### SUDMEX-CONN dataset

##### Participants

In this study, we used the Mexican magnetic resonance imaging dataset (SUDMEX-CONN) ^63^ as a discovery cohort to establish a machine learning-driven FC signature for CUD phenotyping. In SUDMEX-CONN dataset, recruited CUD patients meet the criteria of having used cocaine for at least one year and with an average of at least three times a week, and no more than 60 days of abstinence in the past year ^63^. Before and on the day of the study, any drug intake for the participants was prohibited. Prior to the MRI scanning, participants were either active cocaine users or had been abstinent for less than 60 days. Additional exclusion criteria included: any electronic or metal implants; history of schizophrenia, bipolar disorder, mania, or hypomania; history of any heart-related disease such as myocardial infarction currently, and so on. Participants aged 18 to 50 years, who provided informed consent, were scanned between March 2015 and October 2016. Further information about the subject recruitment criteria can be found in the literature ^63^. Our study focused on 135 participants who have both diagnosis labels and neuroimaging data.

##### Clinical assessments

The diagnosis of cocaine dependence was made using the *MINI International Neuropsychiatric Interview – Plus Spanish version 5.0* (MINI) ^64^. It should be noted that our study did not distinguish cocaine dependent disorder, and cocaine use disorder as the boundaries between these disorders are not clearly defined ^65^. Additionally, diagnoses for alcohol abuse, substance/drug abuse, and suicide risk were also evaluated using MINI. The severity of cocaine craving was assessed by *Cocaine Craving Questionnaire General* (CCQ-G) ^66^. The *World Health Organization Disability Assessment Schedule 2.0* ^67^ (WHODAS) was used to evaluate the functional impairments and disabilities of participants. It consists of six domains: cognition, mobility, self-care, getting along, life activities, and participation. The impulse control of participants was measured using the *Barratt Impulsiveness Scale version 11* (BIS) ^68^ in three domains: cognitive impulsiveness, motor impulsiveness, and non-planning impulsiveness. The psychological symptoms and distress of participants were assessed using the *Symptom Checklist-90- revised* (SCL) ^69^, which covers nine behavioral domains: somatization, obsessive-compulsive disorder, anxiety, depression, interpersonal sensitivity, hostility, paranoid ideation, phobic anxiety, and psychoticism.

##### Resting-state fMRI acquisition and preprocessing

Resting-state fMRI was scanned using gradient echo planar imaging pulse sequences. The scanning parameters were set as follows: repetition time/echo time = 2000/30 ms, flip angle = 75°, matrix = 80 × 80, field of view = 240mm^2^, voxel size = 3 × 3 × 3 mm, number of slices = 36. Participants were instructed to keep their eyes open during the scanning process. The acquired resting-state fMRI data were preprocessed using the reproducible fMRIPrep pipeline ^70^. The T1 weighted image was corrected for intensity nonuniformity and then skull was stripped as T1w reference. Spatial normalization was done through nonlinear registration with the T1w reference ^71^. The brain tissue (cerebrospinal fluid, white matter, and grey matter) was segmented from the T1w reference using FAST (FSL) ^72^. The BOLD reference was then transformed to the T1w reference using a boundary-based registration method, configured with nine degrees of freedom to account for distortion remaining in the BOLD reference ^73^.

Head-motion parameters (six rotation and translation parameters of volume-to-reference transform matrices) were estimated with MCFLIRT (FSL) ^74^. BOLD signals were slice-time corrected and resampled onto the participant’s original space with head-motion parameters, susceptibility distortion correction, and then resampled into standard space, generating a preprocessed BOLD run in MNI152NLin2009cAsym space. Automatic removal of motion artifacts using independent component analysis (ICA-AROMA) ^75^ was performed on the preprocessed BOLD time-signals in MNI space after removal of non-steady-state volumes and spatial smoothing with an isotropic Gaussian kernel of 6 mm FWHM (full-width half-maximum). Six participants with excessive movement (>0.5 mm motion framewise displacement) ^76, 77^ were excluded, leaving 129 subjects with usable fMRI data for analysis. The demographic information of these subjects is summarized in Table S4.

### Replication Cohort

We utilized and harmonized CUD patients from the New York University dataset and SUDMEX-TMS dataset, and healthy controls from the UCLA-CNP dataset to form a replication cohort for independent validation of the identified FC signature. All demographic information of these datasets used for the independent cohort are summarized in Tables S5-S7, respectively. The overall demographic information of the independent cohort is summarized in Table S8.

#### New York University dataset

In this dataset, the study recruited right-handed adults aged 28 to 46 who had a diagnosis of cocaine dependence within the last year according to ‘*Diagnostic and Statistical Manual of Mental Disorders*’ DSM-IV criteria and had been abstinent for at least two weeks. The participants were scanned with fMRI at the New York University School of Medicine and the VA New York Harbor Healthcare System. The exclusion criteria included: positive result of the cocaine test on the scanning day, an incidental brain abnormality, and four exhibited excessive movement ^9^. This dataset consists of 29 CUD patients and 24 healthy controls (http://fcon_1000.projects.nitrc.org/indi/retro/nyuCocaine.html). However, only data from the 29 CUD patients were publicly available and thus used for our analysis.

Resting-state fMRI data were collected using multi-echo EPI sequences from Siemens Allegra 3T in 6 mins. The scanning parameters included: 180 time points; TR = 2000 ms; flip angle = 90°; number of slices = 33; voxel size = 3 × 3 × 4 mm). Participants were scanned with their eyes open.

#### SUDMEX-TMS dataset

This dataset was collected by a clinical trial involving rTMS treatment, conducted at the Clinical Research Division of the National Institute of Psychiatry in Mexico City. All procedures were approved by the Institutional Ethics Research Committee and registered (CEI/C/070/2016; ClinicalTrials.gov NCT02986438). A total of 54 CUD patients (ages 18 – 50) were recruited and met the inclusion criteria such as high cocaine consumption for at least one year ^25^. Baseline resting-state fMRI data of one patient was unpublished online, and hence 53 patients were used for our replication cohort. This dataset was also used in our prediction analysis of rTMS treatment outcome (refer to ‘Association Between Discriminative Connections and TMS treatment response’ subsection in Results). More details involving rTMS treatment could be found in the method section ‘Clinical Trial Cohort with rTMS Treatment’.

Resting-state fMRI data were acquired using a gradient echo-planar pulse sequence of Philips Ingenia 3T scanner with 32-channel head coil. The acquisition parameters of the scanners were listed as followings: TR/TE = 2000/30 ms, FOV = 240 mm, flip angle = 75°, matrix size = 70 × 70, voxel size = 3 × 3 × 3.33 mm, slice thickness = 3.33 mm, number of slices = 37, duration = 10 min).

#### UCLA-CNP dataset

To ensure the reliability of our findings, we constructed a replication cohort for independent validation of FC signature findings by matching the age, gender and size of CUD patients from the New York University dataset and the SUDMEX-TMS dataset. We selected the corresponding healthy controls from the UCLA-CNP dataset to create the replication cohort. Resting-state fMRI data of 81 healthy controls (aged 21 to 50) was utilized ^78^. The discovery cohort and replication cohort have similar age and gender distributions between CUD group and healthy control group (Table S4 and S8).

Resting-state fMRI data were acquired by T2* weighted images using a single-shot gradient echo-planar pulse sequence. They were collected with 3 Tesla scanners lasting for 304 seconds with different scanners but the same parameter set (repetition time = 2000 ms, echo time = 30 ms, flip angle = 90°, matrix size = 64×64, FOV = 192 mm, slice thickness = 4 mm, slices number = 34).

#### Resting-state fMRI preprocessing

The same preprocessing pipeline as used in the discovery cohort was adopted for preprocessing fMRI data in the replication cohort.

#### Clinical Trial Cohort with rTMS Treatment Participants

The SUDMEX-TMS clinical trial dataset was used here again for investigating the association of identified connectivity signature and response to rTMS treatment. Nine of the recruited 54 CUD patients did not receive the allocated intervention, leaving 25 patients in the active rTMS and 20 in the sham rTMS, following a double-blind randomized controlled trial. In the maintenance phase: 20 patients (15 initially assigned to active and 5 to sham) completed 3 months of twice-weekly rTMS sessions; while 15 patients (10 initially assigned to active and 5 to sham) completed 6 months of rTMS sessions, and 7 patients (3 initially assigned to active and 4 to sham) completed 12 months of twice-weekly rTMS sessions. A MagPro R301+Option magnetic stimulator and a figure-of-eight B65-A/P coil (MagVenture, Alpharetta, GA) were used. During the 10 weekdays of the first two weeks, 5000 pulses per day were delivered at 5 Hz. Stimulation was delivered at 100% motor threshold to the left dorsolateral prefrontal cortex. The maintenance phase comprised two 5-Hz (5000 pulses per day) sessions per week. The severity of the cocaine craving was assessed by the difference in craving visual analog scale scores between baseline and after two weeks of treatment. More details of the experiment design can be found in the literature ^25^. The information of resting-state fMRI acquisition and preprocessing can be found in the method section ‘SUDMEX-TMS dataset’. In this study, we focused on the patients in double-blind 2- week rTMS treatment phase, since the sample size in the maintenance phase is limited.

#### Functional Connectivity Calculation

The voxel-level BOLD signals were averaged into 100 regional time series according to the Schaefer parcellation ^29^. The functional connectivity was computed by Pearson’s correlation (Fisher Z-transformed) between every pair of regional time series (Figure S10A). We regressed out framewise displacement from FCs for these cohorts to alleviate the head motion effect (Figure S11).

#### Data Augmentation

To enhance model generalizability and avoid overfitting, data augmentation is a commonly employed technique in machine learning when the available sample size is limited ^79–81^. The discovery cohort comprised 10-min scans of 300 volumes of fMRI for each subject. We subsampled full-length BOLD signals from each subject into two 5-min time series for data augmentation since this data length has been reported to be sufficient for calculating reliable functional connectivity ^82^. The FCs constructed from whole time signals and twice-augmented FCs from two 5-min time series were utilized as the training set. A comparison analysis was conducted to investigate the effects of data augmentation times on training the classification model. We found that twice data augmenting led to improved classification performance that was stable (Figure S12).

#### Classification Model Training and Testing

We employed the XGBoost algorithm, an advanced ensemble learning technique^26^, to classify CUD patients and healthy controls. By leveraging resting-state FC features extracted from our discovery cohort, we built robust machine learning models using this algorithm combined with data augmentation and feature selection. To reduce the number of FC features, two-sample t-tests were used to identify significantly different FCs between CUD patients and healthy controls, using a threshold of p < 0.05.

XGBoost was then trained with the selected FC features to classify subjects as CUD patients or healthy controls. The classification workflow is summarized in Figure S10B. The hyperparameters of XGBoost were set as follows: learning rate = 0.5, maximum depth of a tree = 5, L1 regularization = 10, L2 regularization = 15, and the number of trees = 10. The classification performance was evaluated by 10- fold cross-validations for metrics including accuracy, sensitivity, and specificity. It should be noted that the statistical analysis-based feature selection step was done on the training set alone. The generated feature mask will then be applied to the test set for feature selection and classification.

To further validate the generalizability of the identified FC signature to unseen data, we applied ten well-trained XGBoost models from each run of the cross-validation to predict diagnostic labels of subjects in an independent replication cohort. The predicted labels were determined by averaging the probabilities of predicted labels from the ten models, followed by classification performance evaluation. Site effects in FC features were examined using statistical t-tests between different cohorts (Figure S13). To facilitate the replication analysis, we applied ComBat ^83^, a powerful site-effect correction method, to harmonize the FCs across different datasets based on demographic information including gender and age.

## Data Availability

All data produced are available online at
http://fcon_1000.projects.nitrc.org/indi/retro/nyuCocaine.html
https://openneuro.org/datasets/ds000030/versions/00016
https://openneuro.org/datasets/ds003037/versions/1.0.1
https://openneuro.org/datasets/ds003346/versions/1.1.2

## Acknowledgement

This work was supported in part by NIH grant nos. R01MH129694 and R21MH130956, and Lehigh University FIG (FIGAWD35), CORE, and Accelerator grants. Portions of this research were conducted on Lehigh University’s Research Computing infrastructure partially supported by NSF Award 2019035. A.E. was supported by NIH grant nos. DP1MH116506 and R44MH123373. G.A.F. was supported by NIH grant nos. K23MH114023 and R01MH125886 and grants from the Brain and Behavior Research Foundation and One Mind – Baszucki Brain Research Fund.

## Supplementary Materials

**Figure S1.**
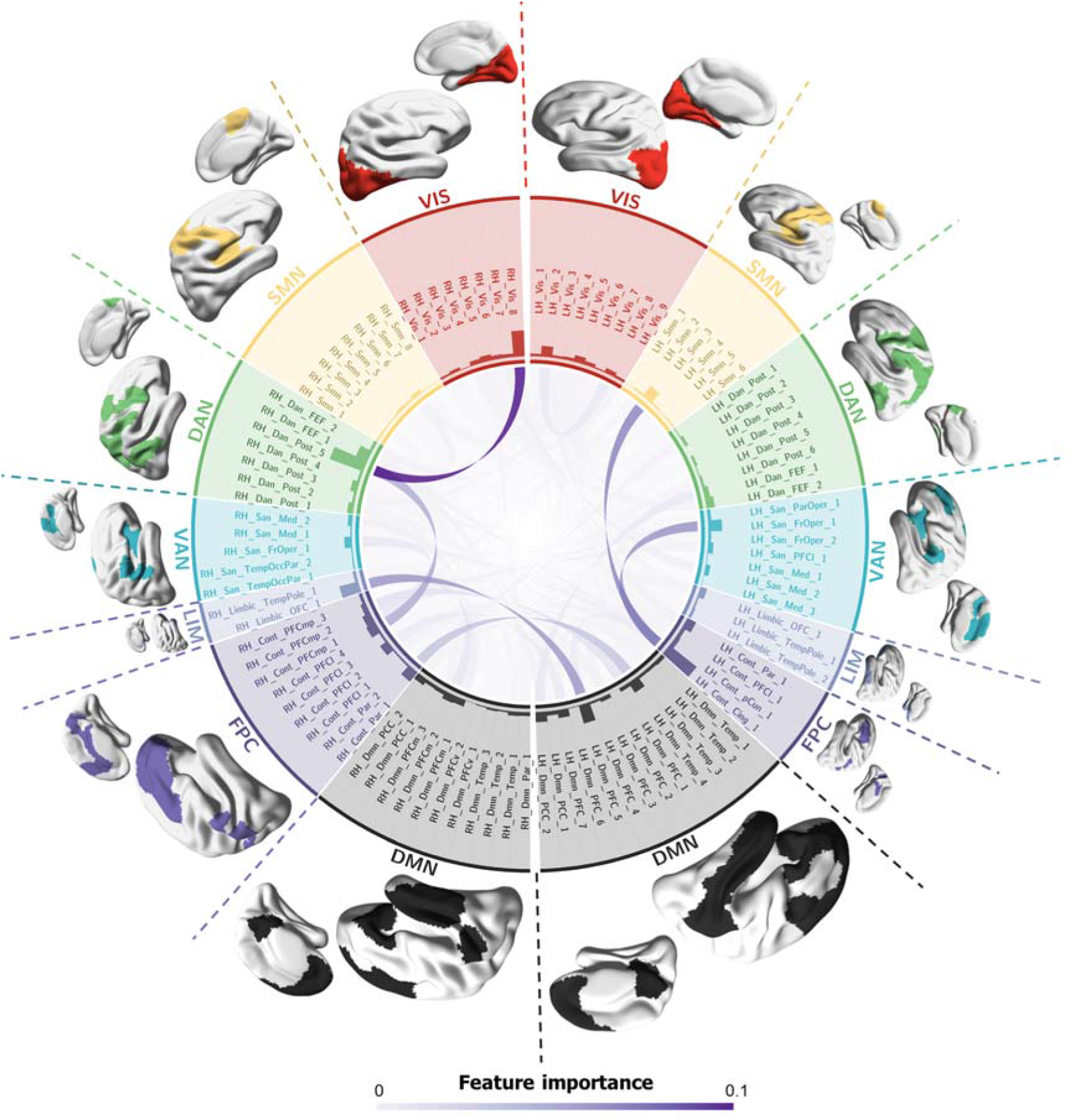
Visualization of all CUD-discriminative FCs. Histogram indicated the node strength calculated from the sum of the linked FC importance. VIS, visual network; SMN, somatomotor network; DAN, dorsal attention network; VAN, ventral attention network; LIM, limbic network; FPC, frontoparietal control network; DMN, default mode network.

**Figure S2.**
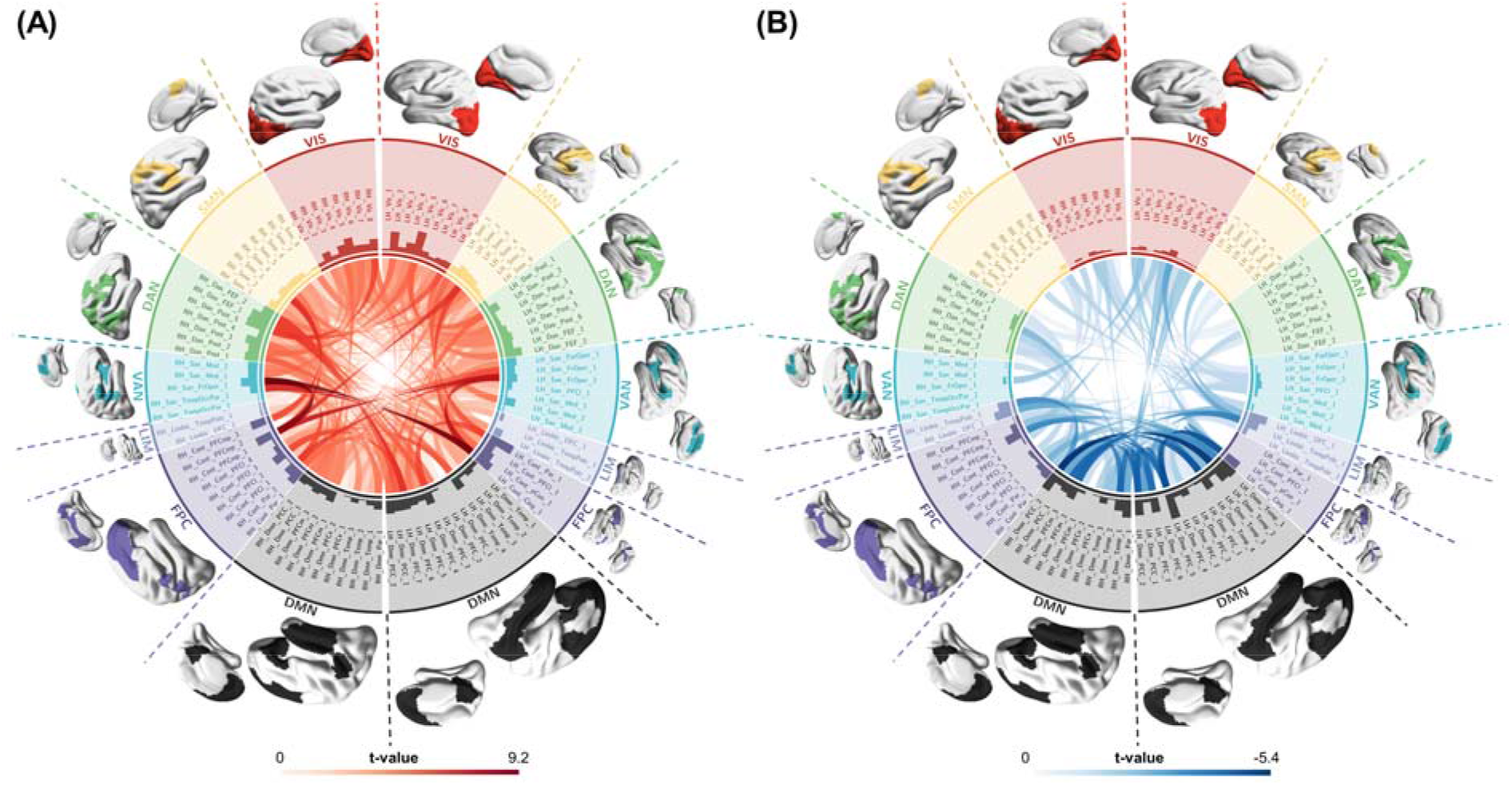
Statistical difference in the classifier-identified discriminative FCs between CUD patients and healthy controls, examined by two-sample t-tests. **(A)** All significant hyperconnections (CUD > healthy controls). **(B)** All significant hypoconnections (CUD < healthy controls). Histogram indicated the node strength calculated from the sum of the linked FC importance. VIS, visual network; SMN, somatomotor network; DAN, dorsal attention network; VAN, ventral attention network; LIM, limbic network; FPC, frontoparietal control network; DMN, default mode network. Only the significant t values that survived FDR were shown.

**Figure S3.**
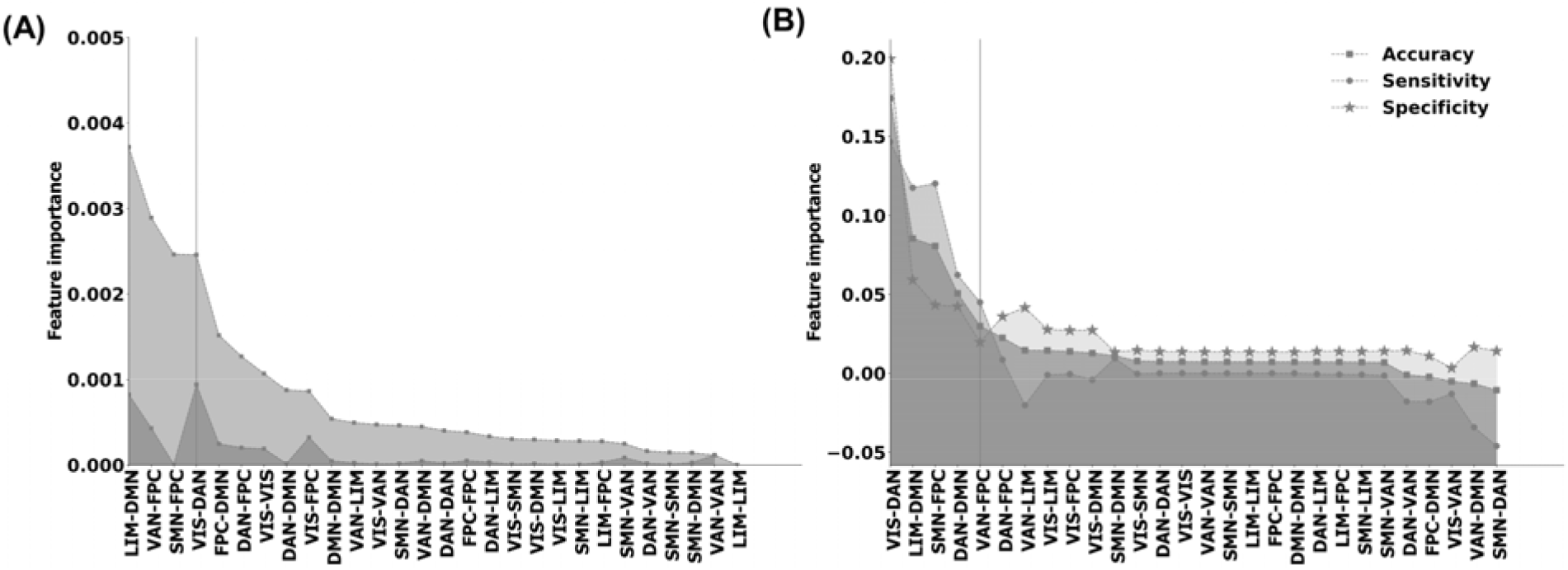
The importance of network-level functional connectivity. **(A)** We grouped the importance of FC features defined from the frequency of the feature occurrence in XGBoost models based on Yeo’s 7 networks. Then the network-level importance sorted in descending order, was shown in light grey. The difference between sorted and adjacent importance was calculated and visualized in deep grey. The network importance is dramatically decreased with the largest adjacent difference when it comes to the VIS-DAN connectivity. **(B)** To further assess the significance of important FCs for further analyses, we randomly permuted FCs in each pairwise network 1000 times and calculated the difference between the average permute performance and the raw performance as feature importance. The sorted importance was plotted. The top 5 important network FCs, which were significantly (p_fdr_ < 0.05) larger than the permuted results, were on the left of the vertical line. Only 4 network-level FCs with significant importance were observed in both calculation strategies (VIS-DAN, LIM-DMN, SMN-FPC, VAN-FPA). Therefore, we only analyzed the association between the top 4 discriminative network-level FCs with cognitive behavioral assessments.

**Figure S4.**
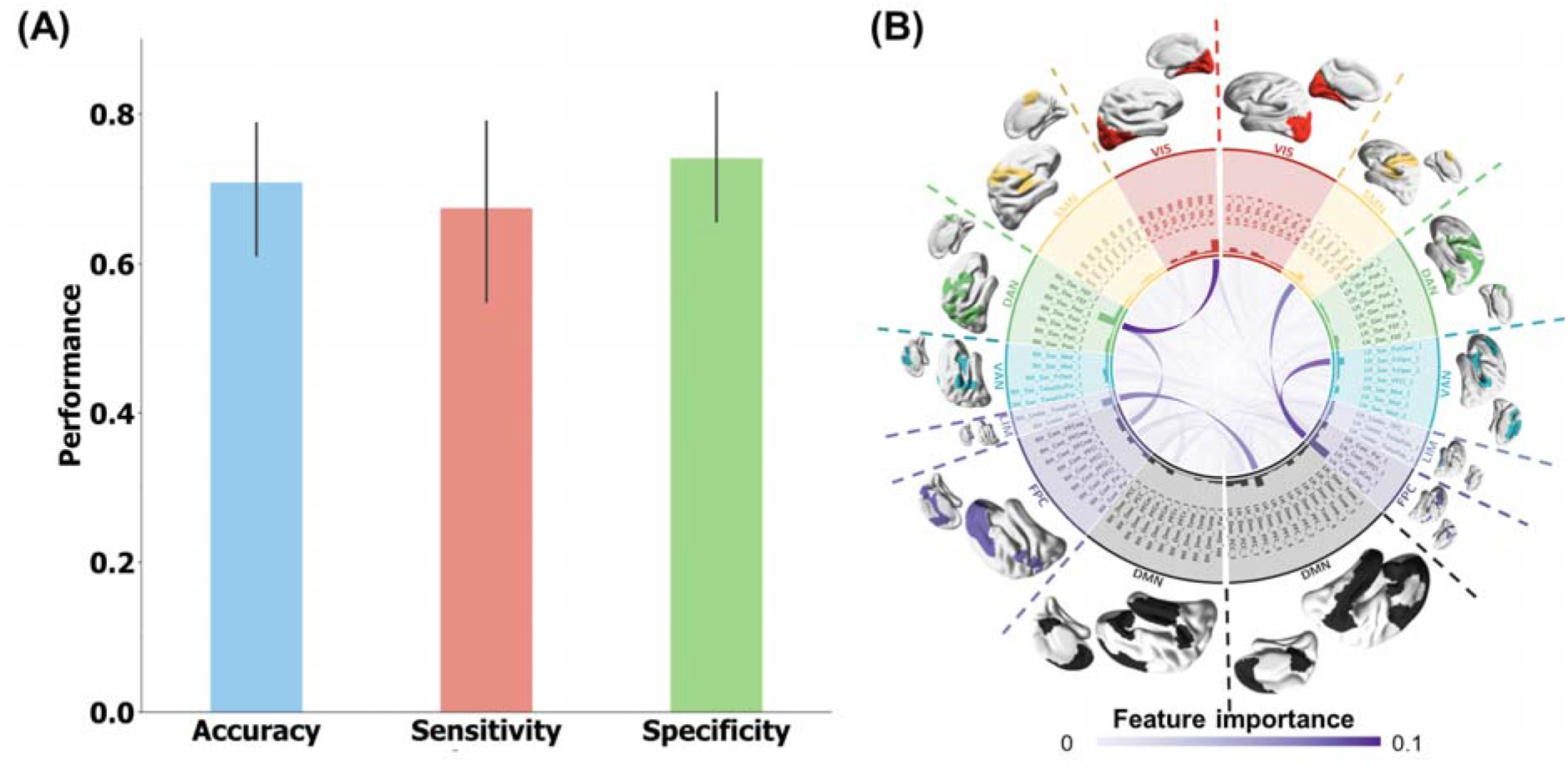
Classification of the CUD and HC in discovery cohort with comorbid confounder control. We regressed out the comorbid variables for each FC, using the simple linear regression models and then trained the models with same hyperparameters in 10-fold cross validations. **(A)** Averaged classification performance: the accuracy, sensitivity, and specificity are 0.71 ± 0.14, 0.67 ± 0.20, and 0.74 ± 0.14, respectively. **(B)** The CUD-discriminative signature.

**Figure S5.**
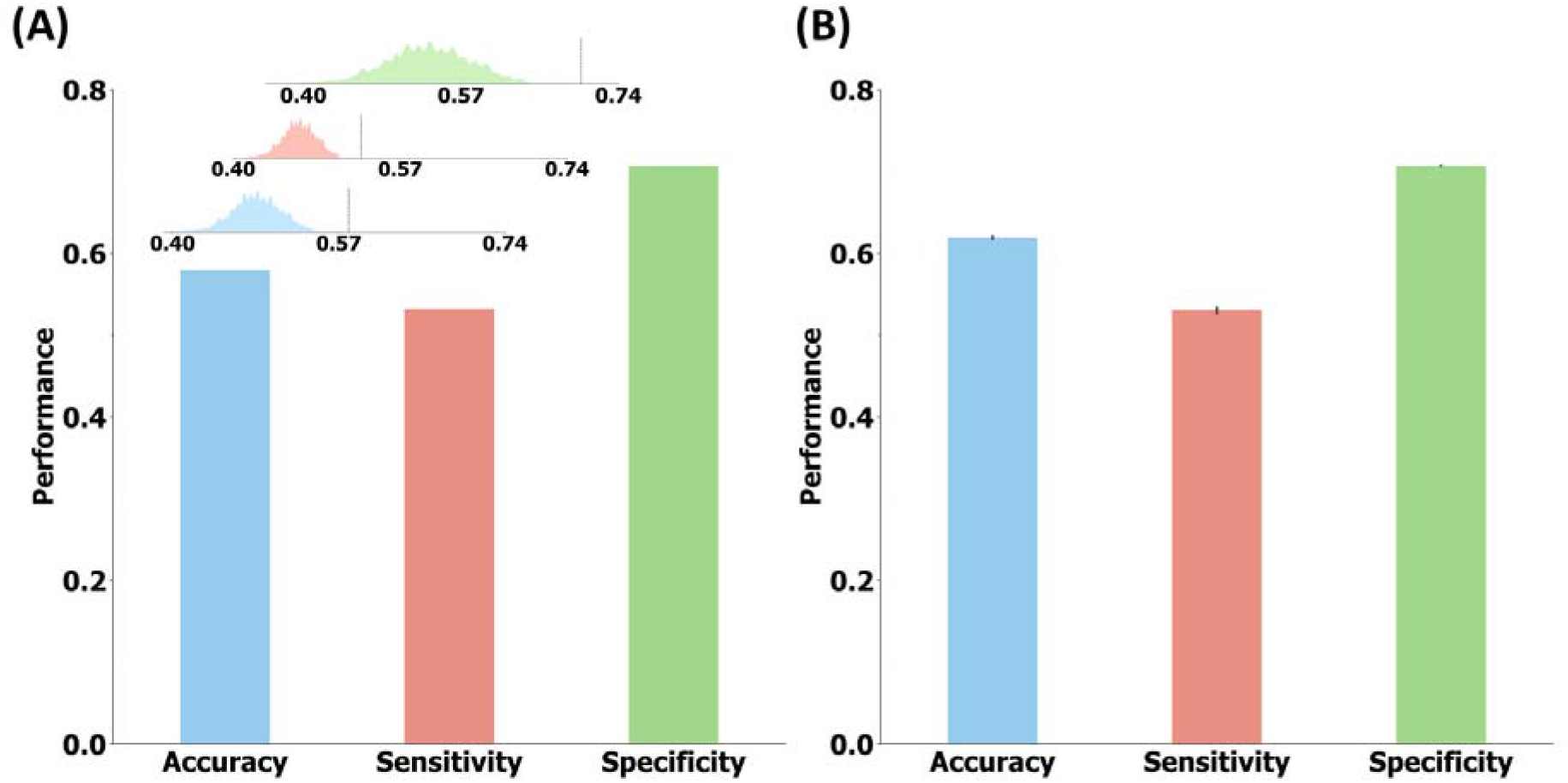
Site classification. All subjects (including 145 patients with multiple mental disorders such as attention-deficit/hyperactivity disorder, bipolar disorder) from UCLA-CNP were used for classification. **(A)** The performance of classifying all subjects from UCLA-CNP or the combination of NYU and SUDMEX-TMS datasets (accuracy = 0.58; sensitivity = 0.53; specificity = 0.70). The performance of distribution was from 1000 random permutations, accuracy_mean_ = 0.48; sensitivity_mean_ = 0.47; specificity_mean_ = 0.54 **(B)** To fairly objectively compare the site classification performance with the reproduced diagnosis classification performance and avoid the size effect of different sample sizes, we randomly subsampling all subjects from UCLA-CNP to the size of healthy controls from UCLA-CNP 1000 times. The average performance randomly subsampling subjects from UCLA-CNP or the combination of NYU and SUDMEX-TMS datasets (accuracy = 0.61 ± 0.03; sensitivity = 0.53 ± 0.06; specificity = 0.70 ± 0) was plotted.

**Figure S6.**
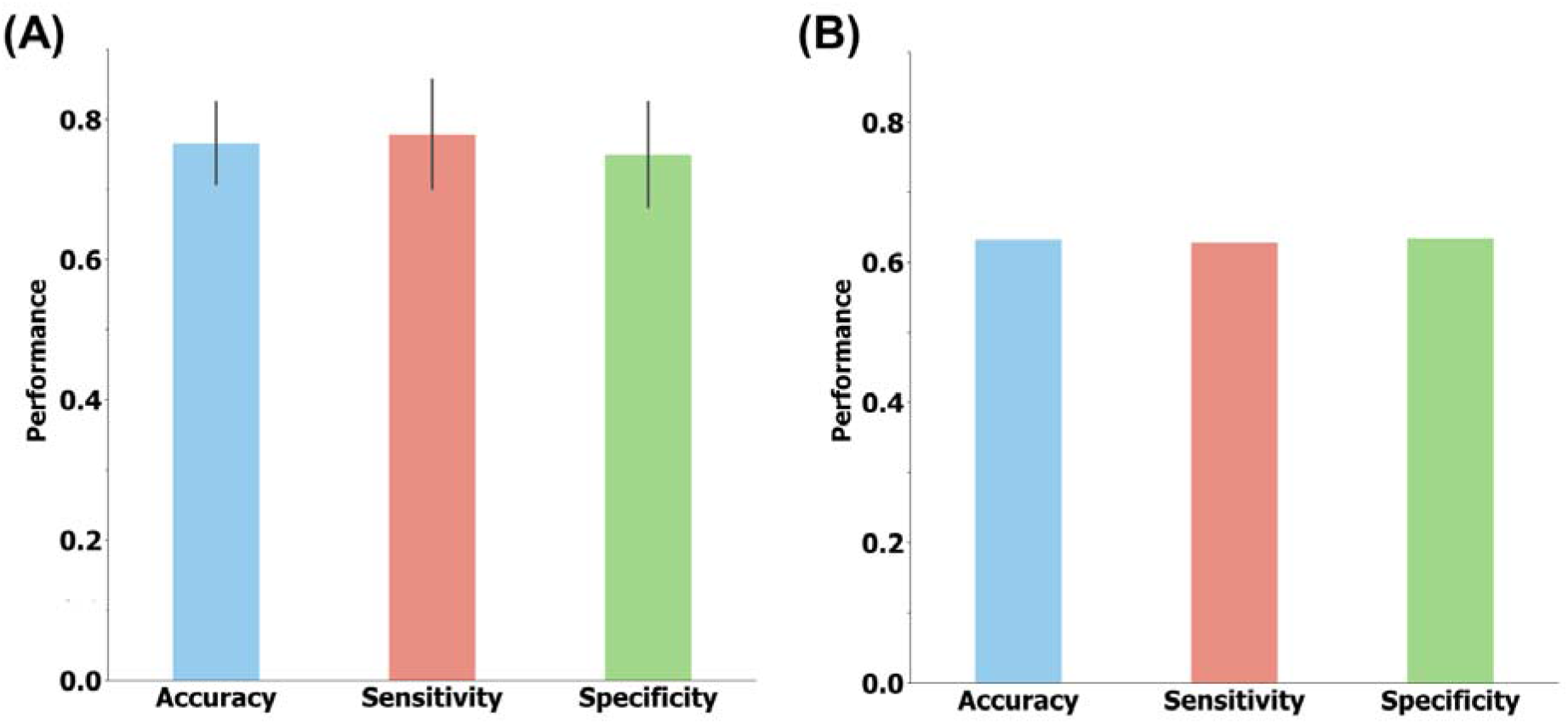
CUD classification reversely. To further verify that the generalizability of our CUD discriminative FC signatures in independent cohort was not primarily dominant by the site effect, we trained the classification models in the independent cohort with same strategy in Figure S5B first. The performance was evaluated with ten-fold cross validation. Then, the obtained discriminative signatures were tested using the discovery cohort. **(A)** The accuracy, sensitivity, and specificity of models in independent cohort are 0.77 ± 0.10, 0.78 ± 0.12, and 0.75 ± 0.13, respectively. **(B)** The accuracy, sensitivity, and specificity of models applied in the discovery cohort are 0.63, 0.63, 0.63, respectively.

**Figure S7.**
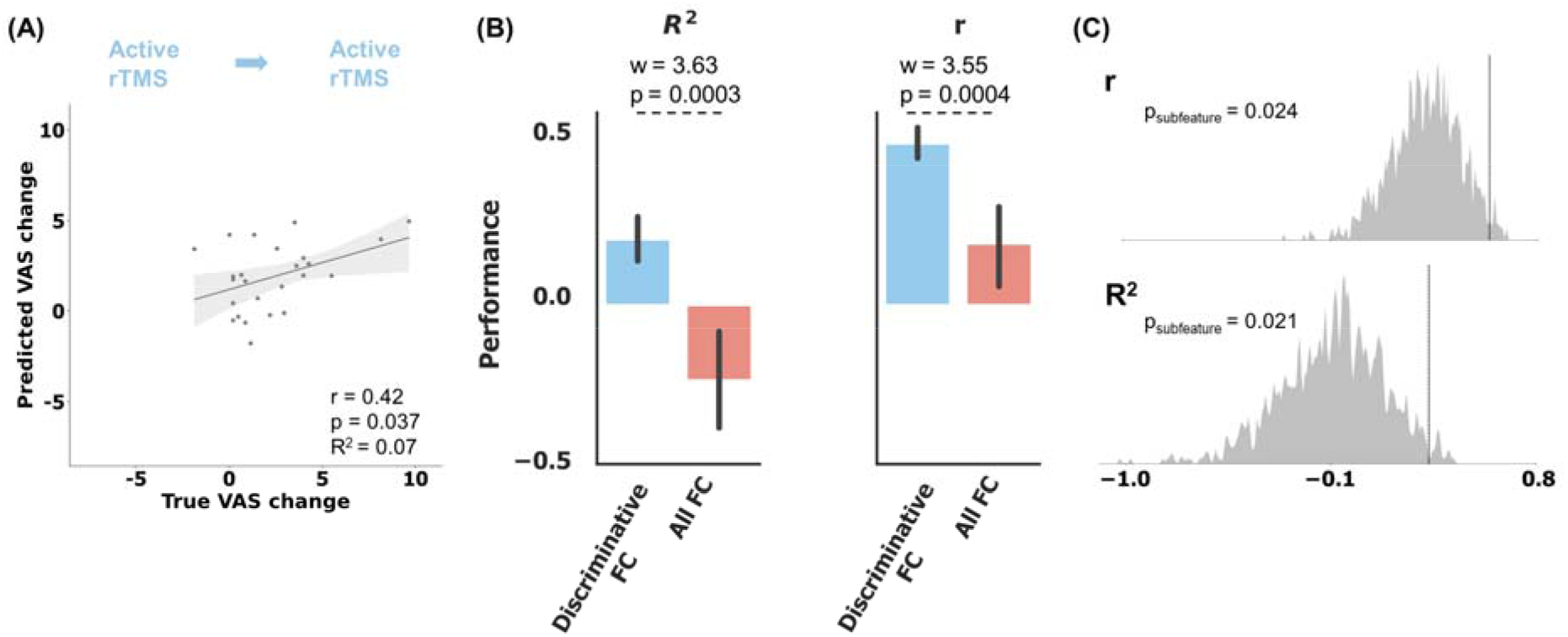
Control study of prediction of the craving visual analog scale (VAS) score changes specific to active rTMS treatment. To verify our hypothesis that the phenotyping FCs own advantage in reflecting the rTMS treatment response, we implement two control studies. **(A)** We used all FCs to predict the active rTMS VAS score change in ten 5-fold cross validations. The predicted scores were averaged from ten models. R^2^ = 0.07 and Pearson’s r = 0.42, P = 0.037 based on the one-sided test against the alternative hypothesis that r > 0. **(B)** The Wilcoxon signed-rank test of the predicted performance from ten models, training with discriminative FC and all FC. The higher R^2^ and r in scatterplot than the mean of ten models were due to the ensemble learning effect ^1^. **(C)** We randomly subsampled all FC features as equal to the size of phenotyping FCs in differentiating CUD and HC. The distributions of the r and R^2^ were shown. The vertical line was the performance from discriminative FC.

**Figure S8.**
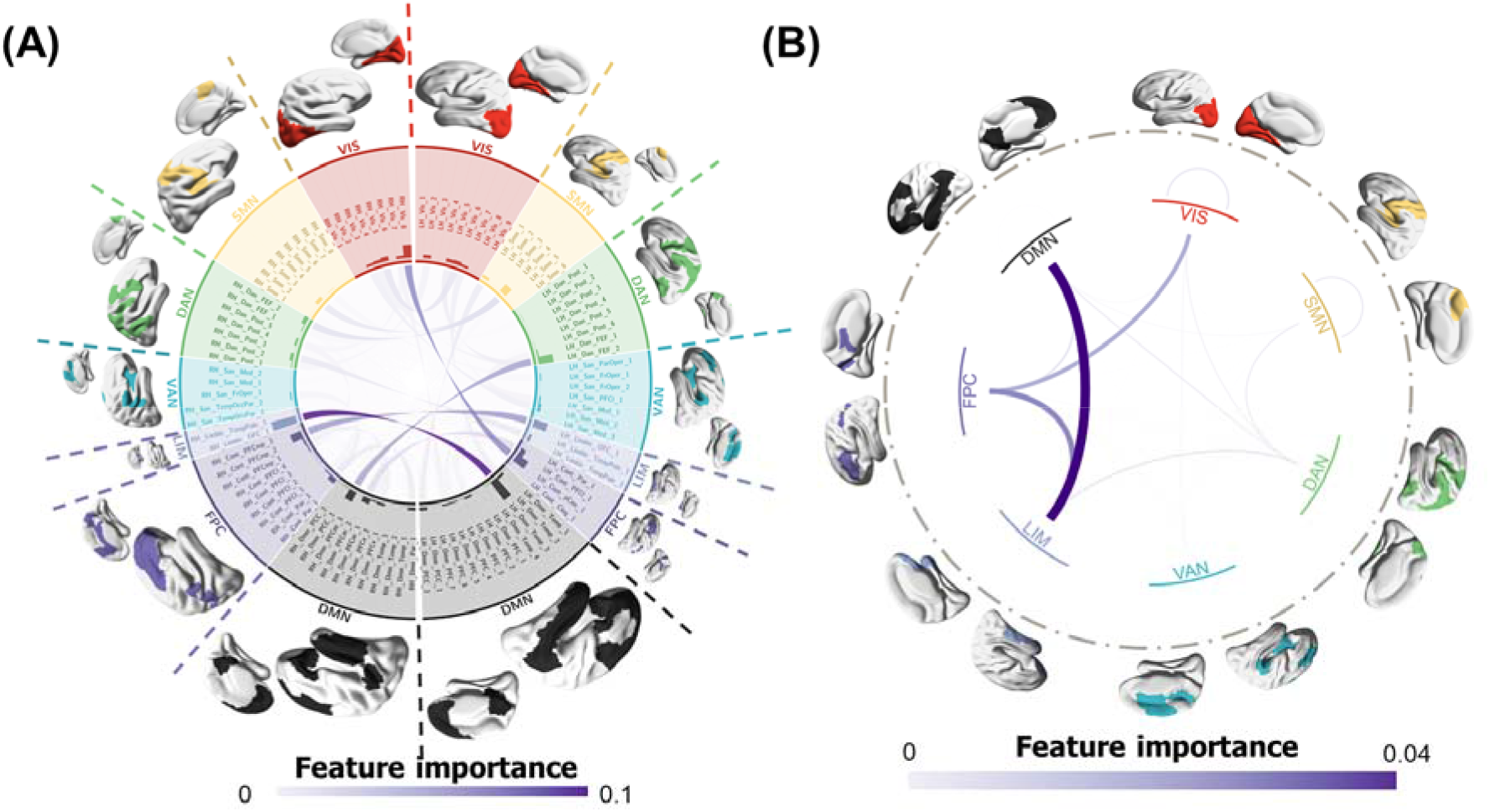
Visualization of the CUD-discriminative FCs involved in repetitive transcranial magnetic stimulation treatment response prediction. **(A)** The rTMS predictive FC signature. **(B)** We grouped the importance of predictive FCs into the seven typical networks including visual network (VIS), somatomotor network (SMN), dorsal attention network (DAN), ventral attention network (VAN), limbic network (LIM), frontoparietal control network (FPC), and default mode network (DMN).

**Figure S9.**
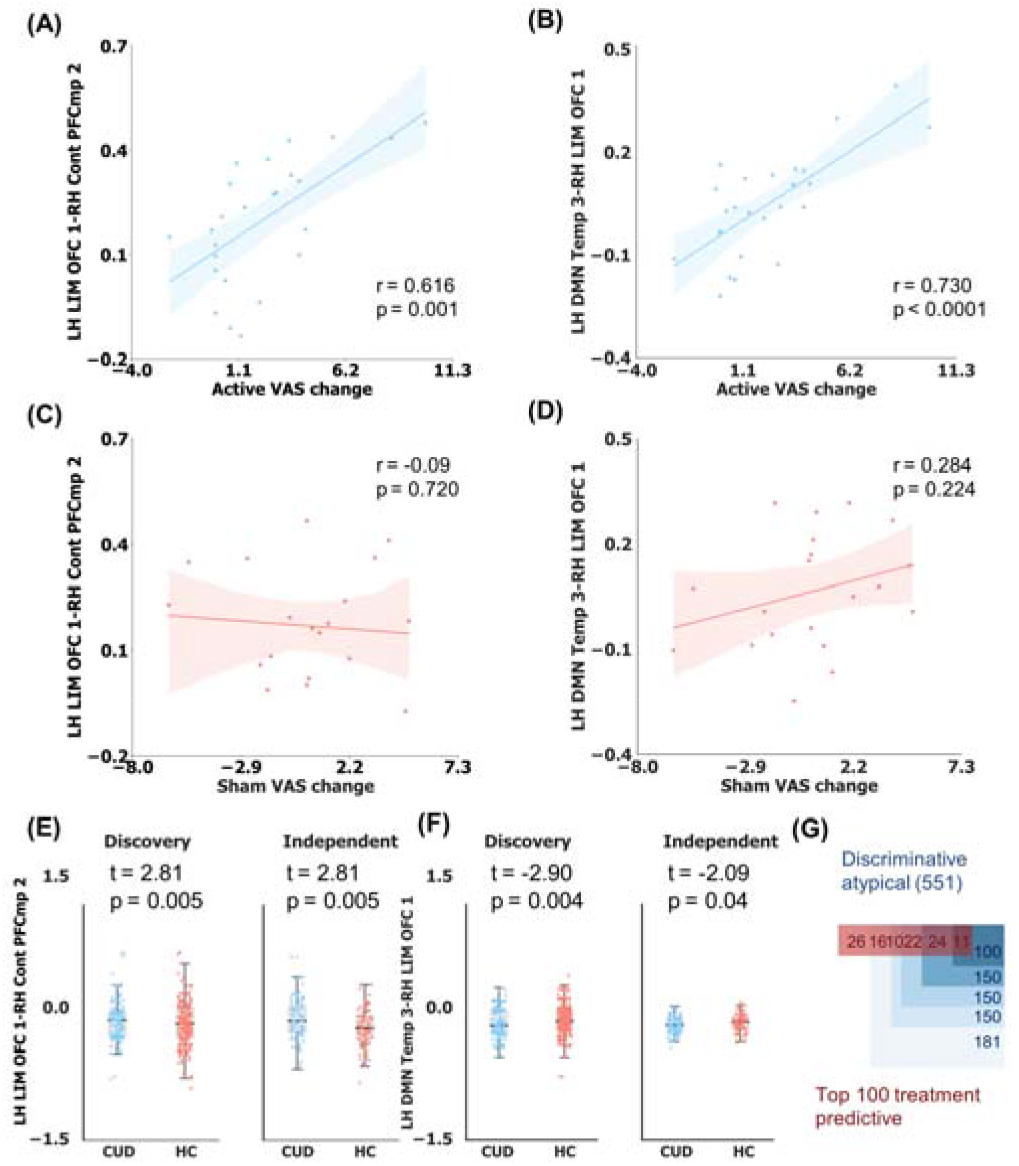
Association between the discriminative FCs and treatment-outcome-predictive FCs. (A), (B) We visualized the correlation between the top 2 active rTMS treatment response predictive FCs and active rTMS VAS score change in scatter plot as illustrative examples. These two FCs were between the oribitofrontal cortex (LH LIM OFC 1) and anterior cingulate cortex (RH Cont PFCmp 2), and between middle temporal cortex (LH DMN Temp 3) and superior oribitofrontal cortex (RH LIM OFC 1). **(C), (D)** The correlation between these two FCs and sham rTMS VAS score change. **(E), (F)** These two FCs distribution between CUD and HC in the discovery and independent cohorts. The data in discovery cohort was augmented twice. These two FCs were significantly and specifically correlated to the VAS score change and significantly different between CUD and HC. **(G)** Venn diagram indicating the association between discriminative and abnormal FCs (551) with active rTMS treatment outcome. Discriminative atypical FCs were defined as the discriminative FCs identified by our classification models and the significantly atypical FCs detected by two-sample t-tests comparing CUD and HC subjects, with those surviving FDR correction (p_fdr_ < 0.05). The number of discriminative atypical FCs was equal to the sum of hyperconnections and hypoconnections. Deeper bluer shading indicates larger treatment predictive weights. The red numbers in the red rectangle represents the overlapping numbers between the top 100 treatment predictive FCs and all discriminative atypical FCs in descending order.

**Figure S10.**
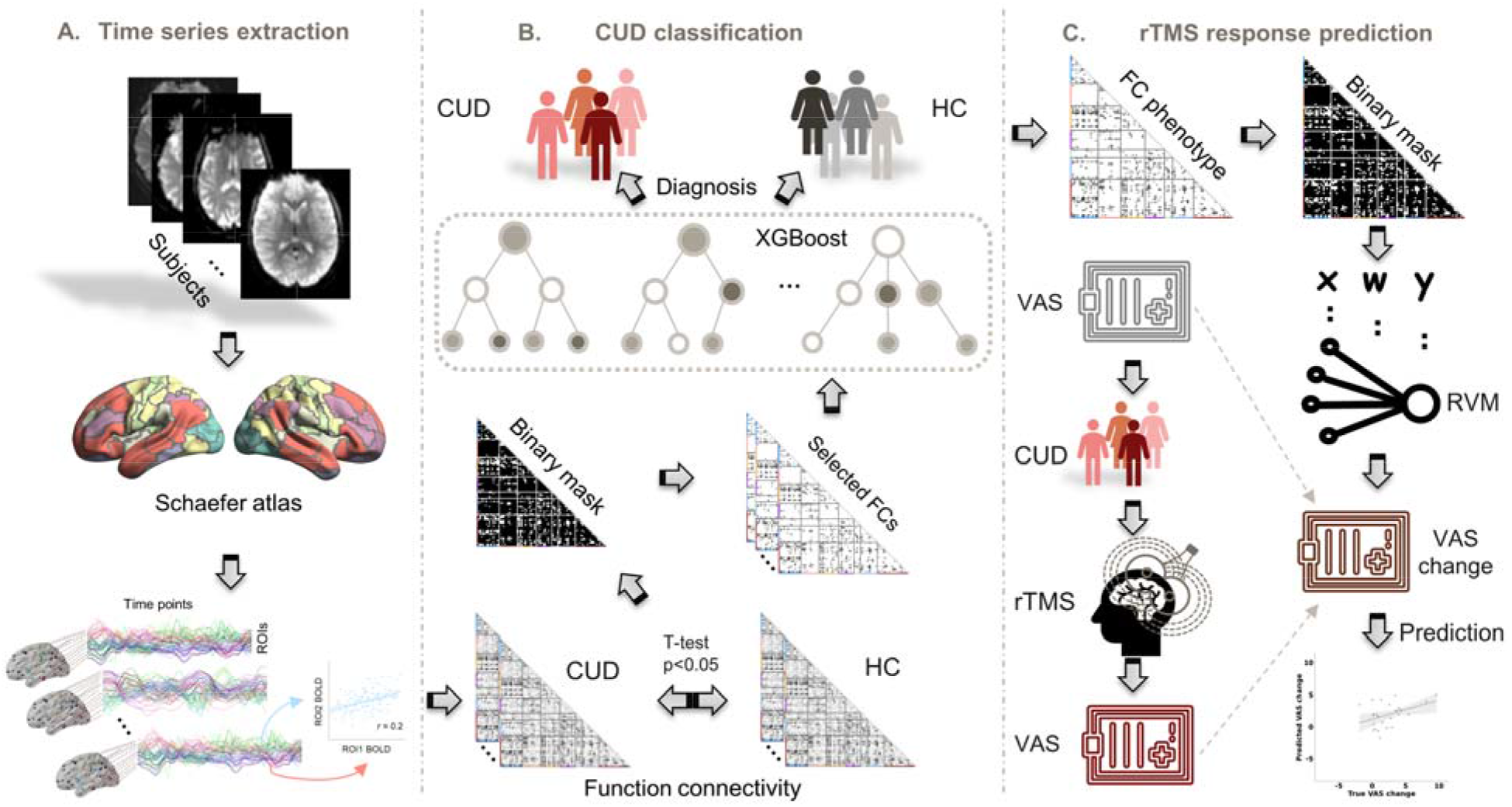
Illustration of our proposed analytical framework. **(A)** Region of interests (ROIs) level time series were extracted from fMRI BOLD signals based on the Schaefer atlas. Functional connectivity was calculated by Pearson’s correlation in time series between any pair of ROIs. **(B)** The functional connectivity features were used to train the XGBoost model to classify the subjects into CUD patients or healthy controls. **(C)** Utilized phenotyping functional connectivity (FC) features, a relevance vector machine (RVM) model was employed to predict changes in visual analog scale (VAS) scores for patients undergoing repetitive transcranial magnetic stimulation (rTMS) treatment.

**Figure S11.**
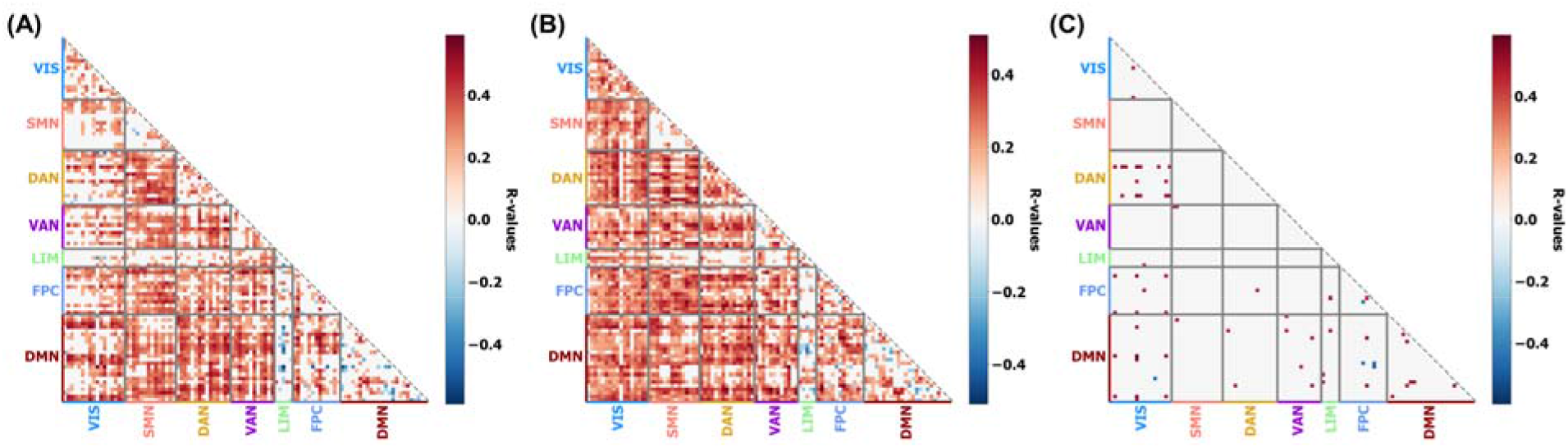
The significant (p_fdr_ < 0.05) Pearson’s correlation between each FC and mean of framewise displacements after FDR correction for each dataset. **(A)** Correlation matrix of the SUDMEX-CONN dataset. **(B)** Correlation matrix of the UCLA-CNP dataset. **(C)** Correlation matrix of the SUDMEX-TMS dataset. Correlation matrix of the NYU dataset is not visualized here since only two FCs significantly correlated to FCs.

**Figure S12.**
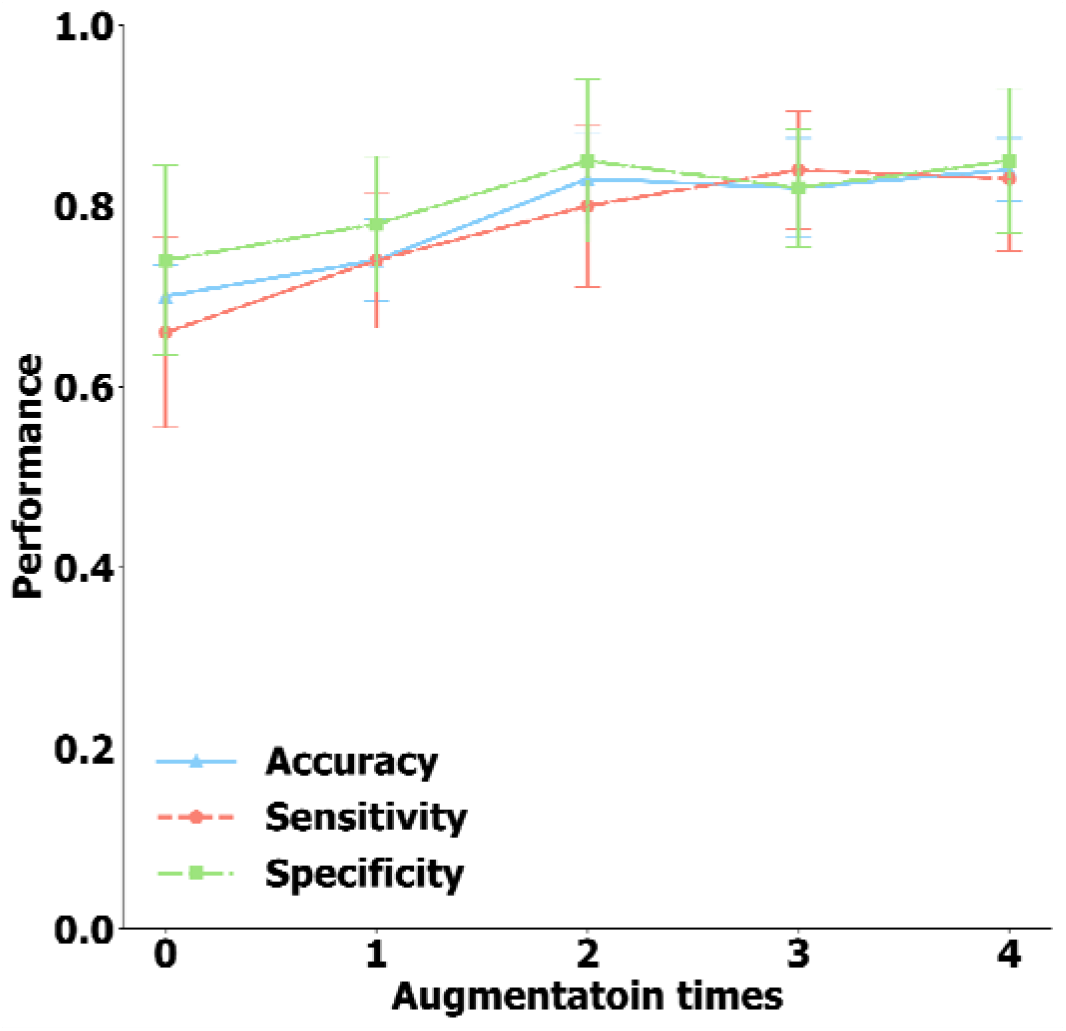
Effects of data augmentation on classification performance. When the augmentation time was equal to 0, the models were trained with FCs extracted from all-sequence time series. When the augmentation time was equal to 1, the models were trained with the FCs extracted from all-sequence time series and from the first semi-sequence time series. When the augmentation time was equal to 2, the models were trained with the FCs extracted from all-sequence time series, from the first semi-sequence time series (the first 150 volumes) and from the second semi-sequence time series (the last 150 volumes). When the augmentation time was equal to 3 or 4, one or two more FC features were extracted from randomly segmented time series with a length of 150 volumes. Without augmentation, the accuracy was 0.70 ± 0.07, sensitivity was 0.66 ± 0.21, and specificity was 0.74 ± 0.16. Applying augmentation once, accuracy was 0.76 ± 0.09, sensitivity was 0.74 ± 0.15, and specificity was 0.78 ± 0.10. Applying augmentation twice, the accuracy was 0.83 ± 0.10, sensitivity was 0.80 ± 0.18, and specificity was 0.85 ± 0.10. Applying augmentation three times, the accuracy was 0.82 ± 0.11, sensitivity was 0.84 ± 0.13, and specificity was 0.82 ± 0.15. Applying augmentation four times, the accuracy was 0.84 ± 0.07, sensitivity was 0.83 ± 0.16, and specificity was 0.85 ± 0.14. Finally, we augmented the FC twice to pursue exhaustive analysis since the performance was no longer further increased.

**Figure S13.**
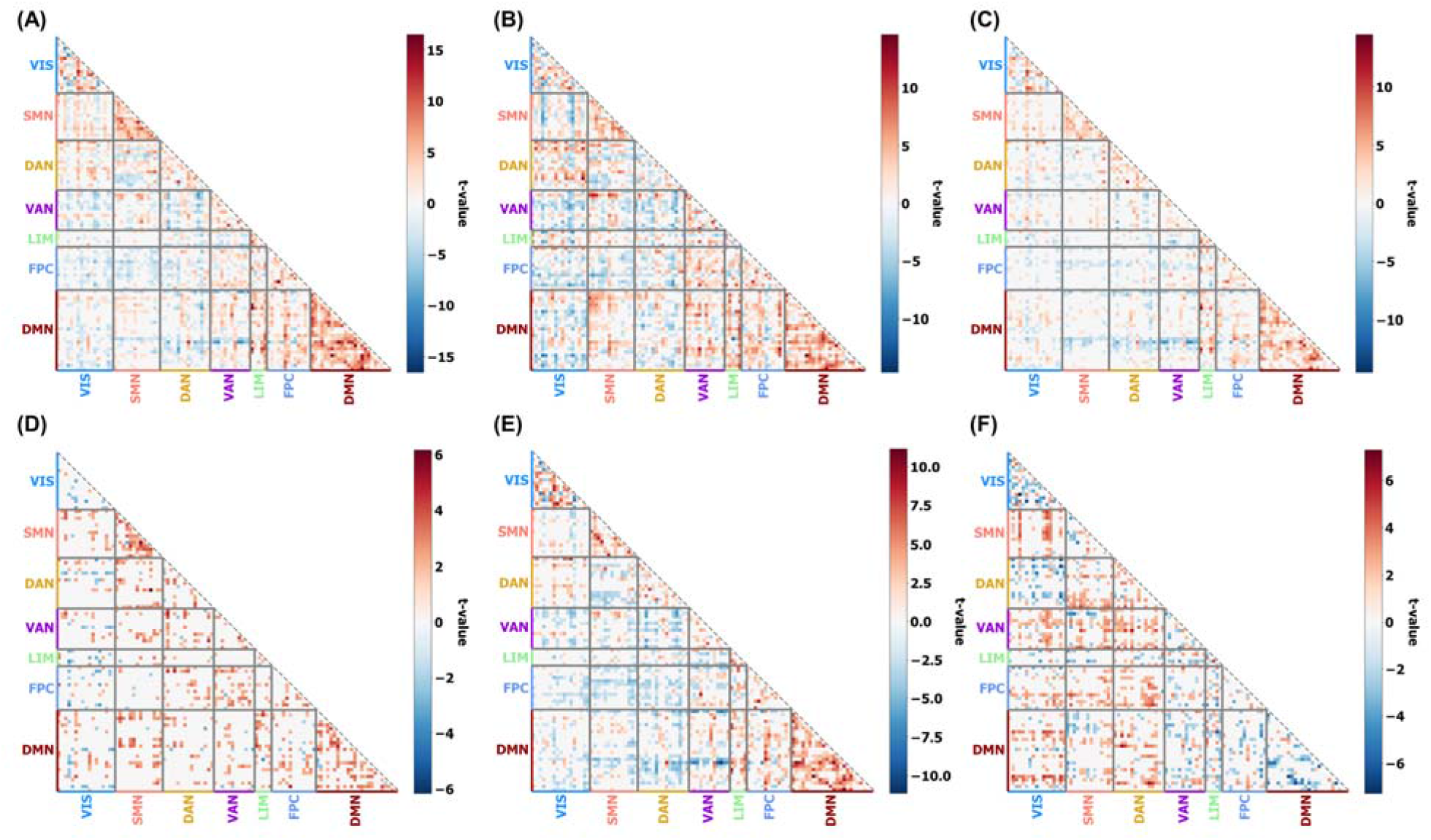
The two-sample t-test comparisons of each FCs across different datasets. **(A)** Comparisons of FCs in SUDMEX-CONN dataset and NYU dataset (SUDMEX-CONN versus NYU). **(B)** Comparisons of FCs in the SUDMEX-CONN dataset and UCLA-CNP dataset (SUDMEX-CONN versus UCLA-CNP). **(C)** Comparisons of FCs in UCLA-CNP dataset and NYU dataset (UCLA-CNP versus us NYU). **(D)** Comparisons of FCs in SUDMEX-CONN dataset and SUDMEX-TMS dataset (SUDMEX-CONN versus us SUDMEX-TMS). **(E)** Comparisons of FCs in SUDMEX-TMS dataset and NYU dataset (SUDMEX-TMS versus us NYU). **(F)** Comparisons of FCs in UCLA-CNP dataset and SUDMEX-TMS dataset (UCLA-CNP versus us SUDMEX-TMS). All t values shown in the panels survived FDR correction (p_fdr_<0.05).

**Table S1.** Results of two-sample t-test comparison of the top 4 discriminative network-level connections between healthy controls and patients diagnosed with other clinical diagnostic labels in MINI International Neuropsychiatric Interview – Plus Spanish version 5.0 (MINI), including substance abuse and suicide diagnosis. These conditions have been suggested to be highly related to CUD 2. The p-values between each connection and all clinical variables (including MINI and the measurements using in Table S2) were corrected for FDR. Significant results from the t-tests, prior to FDR correction (p < 0.05), are displayed in bold and those that passed FDR correction (p < 0.05) are highlighted in italic.

|  | DMN-LIM |  |  | VAN-FPC |  |  | SMN-FPC |  |  | VIS-DAN |  |  |
| --- | --- | --- | --- | --- | --- | --- | --- | --- | --- | --- | --- | --- |
|  | t | p | P<sub>fdr</sub> | t | p | P<sub>fdr</sub> | t | p | P<sub>fdr</sub> | t | p | P<sub>fdr</sub> |
| Suicide risk | **-2.75** | **0.007** | NS | -0.23 | 0.818 | NS | 1.20 | 0.234 | NS | 0.22 | 0.828 | NS |
| Alcohol abuse | **-2.80** | **0.006** | NS | 1.67 | 0.098 | NS | **2.50** | **0.014** | NS | 1.76 | 0.082 | NS |
| Stimulants used history | 0.79 | 0.431 | NS | -0.03 | 0.978 | NS | -0.04 | 0.965 | NS | **-3.36** | **0.001** | **0.011** |
| Hallucinogens used history | **-3.43** | **0.002** | **0.025** | 0.97 | 0.336 | NS | -0.28 | 0.776 | NS | -0.45 | 0.655 | NS |
| Inhalants used history | **-4.43** | **<0.001** | **0.002** | 1.66 | 0.099 | NS | 0.85 | 0.402 | NS | 2.36 | 0.020 | NS |

**Table S2.** Correlation between the top four discriminative network connections and clinical assessments. The scales using here were summarized as follows: World Health Organization Disability Assessment Schedule 2.0 (WHODAS), Cocaine Craving Questionnaire General (CCQ-G), Barratt Impulsiveness Scale version 11 (BIS), Symptom Checklist-90-revised (SCL). Significant results from the t-tests, prior to FDR correction (p < 0.05), are displayed in bold and those that passed FDR correction (p < 0.05) are highlighted in italic.

|  | DMN-LIM |  |  | VAN-FPC |  |  | SMN-FPC |  |  | VIS-DAN |  |  |
| --- | --- | --- | --- | --- | --- | --- | --- | --- | --- | --- | --- | --- |
|  | r | p | P<sub>fdr</sub> | r | p | P<sub>fdr</sub> | r | p | P<sub>fdr</sub> | r | p | P<sub>fdr</sub> |
| WHODAS total | **-0.19** | **0.026** | NS | **0.18** | **0.034** | NS | 0.08 | 0.364 | NS | **0.18** | **0.035** | NS |
| WHODAS cognition | **-0.17** | **0.046** | NS | **0.20** | **0.016** | NS | 0.07 | 0.394 | NS | **0.17** | **0.045** | NS |
| WHODAS mobility | -0.10 | 0.259 | NS | 0.11 | 0.201 | NS | 0.10 | 0.229 | NS | 0.07 | 0.414 | NS |
| WHODAS self-care | -0.03 | 0.740 | NS | 0.15 | 0.075 | NS | 0.12 | 0.175 | NS | 0.04 | 0.674 | NS |
| WHODAS getting along | -0.16 | 0.064 | NS | 0.12 | 0.156 | NS | 0.11 | 0.201 | NS | 0.08 | 0.356 | NS |
| WHODAS life activities | -0.11 | 0.216 | NS | 0.15 | 0.081 | NS | 0.06 | 0.475 | NS | 0.06 | 0.481 | NS |
| WHODAS participation | -0.15 | 0.084 | NS | 0.14 | 0.112 | NS | 0.02 | 0.838 | NS | 0.15 | 0.074 | NS |
| CCQ-G | -0.21 | 0.082 | NS | -0.04 | 0.737 | NS | **-0.26** | **0.031** | NS | 0.06 | 0.593 | NS |
| BIS cognitive | -0.03 | 0.760 | NS | 0.09 | 0.369 | NS | 0.15 | 0.111 | NS | 0.13 | 0.162 | NS |
| BIS motor | -0.07 | 0.462 | NS | 0.13 | 0.168 | NS | 0.14 | 0.143 | NS | 0.14 | 0.145 | NS |
| BIS non-planning | -0.17 | 0.071 | NS | 0.17 | 0.080 | NS | 0.11 | 0.247 | NS | **0.27** | **0.004** | **0.026** |
| BIS total | -0.12 | 0.230 | NS | 0.15 | 0.108 | NS | 0.16 | 0.093 | NS | **0.22** | **0.021** | NS |
| SCL somatization | -0.04 | 0.635 | NS | 0.11 | 0.249 | NS | **0.21** | **0.023** | NS | 0.17 | 0.06 | NS |
| SCL obsessive compulsive | -0.08 | 0.366 | NS | **0.20** | **0.031** | NS | 0.17 | 0.055 | NS | 0.14 | 0.136 | NS |
| SCL interpersonal sensitivity | -0.10 | 0.294 | NS | 0.17 | 0.065 | NS | **0.18** | **0.041** | NS | <0.01 | 0.98 | NS |
| SCL depression | -0.16 | 0.085 | NS | **0.24** | **0.007** | NS | **0.25** | **0.006** | **0.046** | 0.10 | 0.296 | NS |
| SCL anxiety | **-0.19** | **0.041** | NS | **0.23** | **0.011** | NS | **0.26** | **0.004** | **0.038** | 0.03 | 0.706 | NS |
| SCL hostility | -0.08 | 0.399 | NS | **0.21** | **0.021** | NS | **0.21** | **0.018** | NS | 0.13 | 0.158 | NS |
| SCL phobic anxiety | -0.17 | 0.064 | NS | **0.18** | **0.046** | NS | **0.22** | **0.016** | NS | 0.10 | 0.269 | NS |
| SCL paranoid ideation | -0.03 | 0.776 | NS | 0.14 | 0.122 | NS | 0.18 | 0.052 | NS | <0.01 | 0.975 | NS |
| SCL<br>psychoticism | -0.17 | 0.060 | NS | **0.20** | **0.027** | NS | **0.18** | **0.045** | NS | 0.03 | 0.775 | NS |
| SCL total | -0.14 | 0.113 | NS | **0.21** | **0.019** | NS | **0.22** | **0.013** | NS | 0.11 | 0.211 | NS |

**Table S3.** Correlation between the top four discriminative network connections and tobacco use history.

|  | DMN-LIM |  |  | VAN-FPC |  |  | SMN-FPC |  |  | VIS-DAN |  |  |
| --- | --- | --- | --- | --- | --- | --- | --- | --- | --- | --- | --- | --- |
|  | t/r | p | P<sub>fdr</sub> | t/r | p | P<sub>fdr</sub> | t/r | p | P<sub>fdr</sub> | t/r | p | P<sub>fdr</sub> |
| Tobacco use in the last year | **2.08** | **0.040** | NS | **-2.04** | **0.044** | NS | -0.64 | 0.521 | NS | **-2.71** | **0.008** | **0.031** |
| Amount of cigarettes per day | 2.83 | 0.063 | NS | 2.46 | 0.09 | NS | 0.58 | 0.562 | NS | 2.81 | 0.064 | NS |
| Years of tobacco use | **-0.25** | **0.008** | **0.033** | **0.21** | **0.027** | NS | <0.01 | 0.99 | NS | **0.19** | **0.045** | NS |
| Tobacco age of onset | -0.17 | 0.087 | NS | 0.06 | 0.578 | NS | <0.01 | 0.98 | NS | 0.122 | 0.231 | NS |

**Table S4.** Demographic information of the discovery cohort (SUDMEX-CONN dataset).

| Demographic variables |  | HC |  | CUD |  | Statistic values |  |
| --- | --- | --- | --- | --- | --- | --- | --- |
| | | n | % | n | % | $\chi^2$ | p |
| Gender | Male | 47 | 84 | 62 | 87 | 0 | 1 |
|  | Female | 9 | 16 | 9 | 13 |  |  |
|  | unknown | 2 |  | 0 |  |  |  |
|  |  | mean | std | mean | std | t | p |
| Age | | 31.4 | $\pm 8.2$ | 31.2 | $\pm 7.3$ | 0.15 | 0.88 |

**Table S5.**
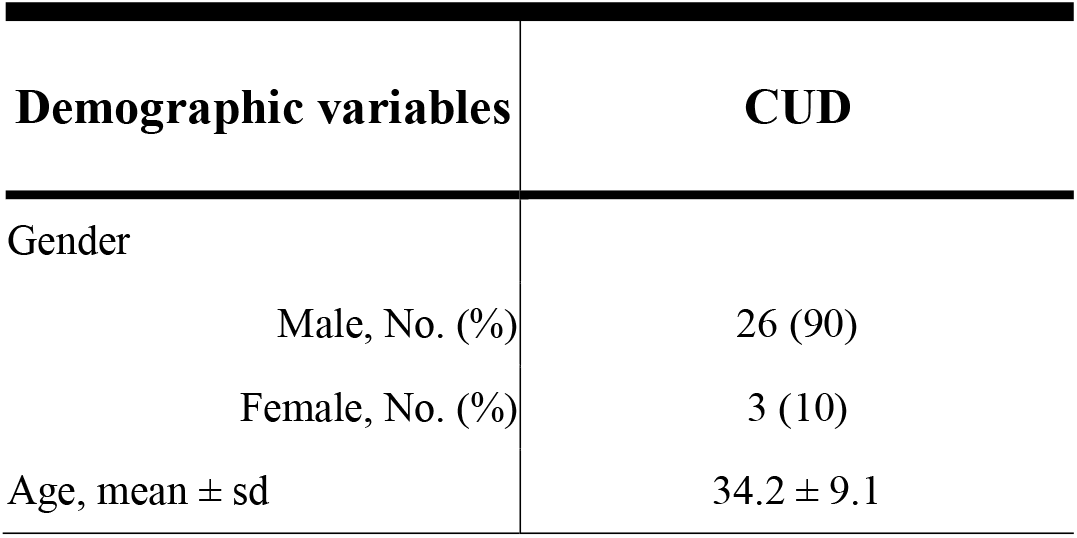
Demographic information of New York University datasets.

**Table S6.**
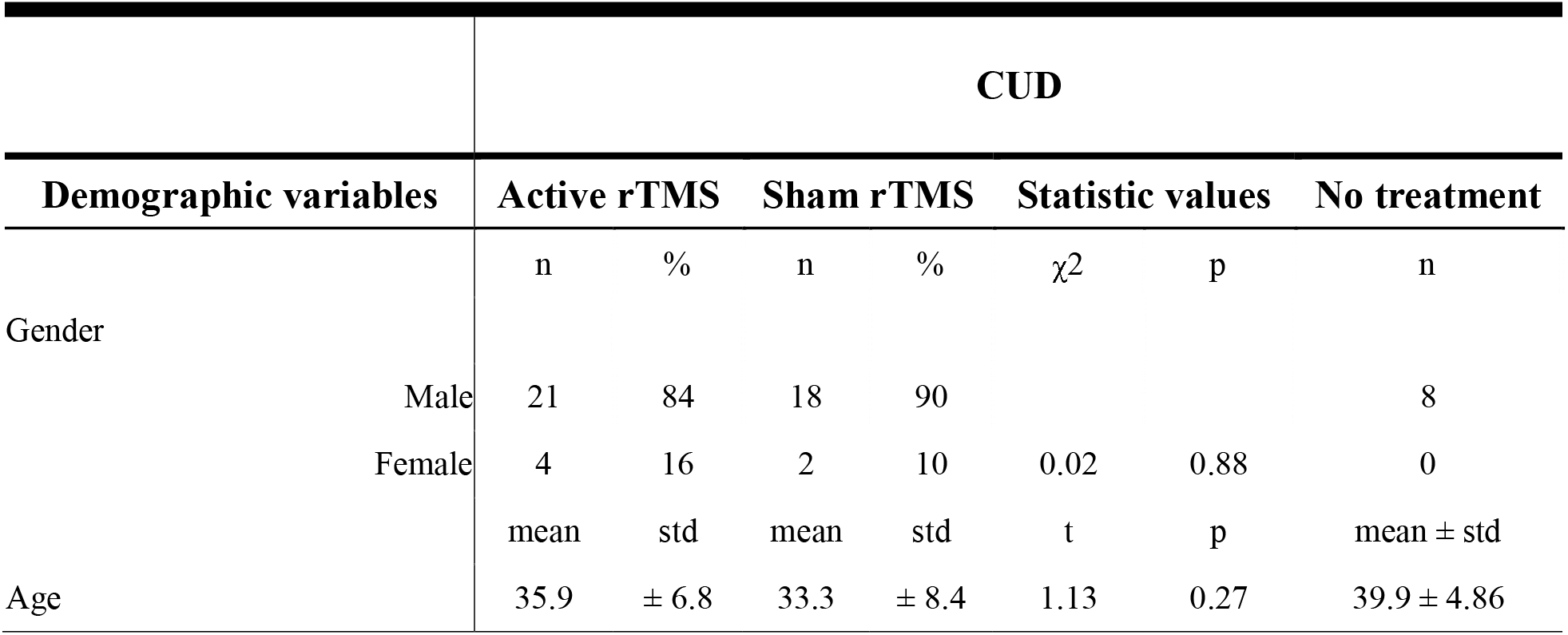
Demographic information of SUDMEX-TMS datasets. The statistical comparison was only applied for the patients with rTMS treatments in two weeks.

**Table S7.**
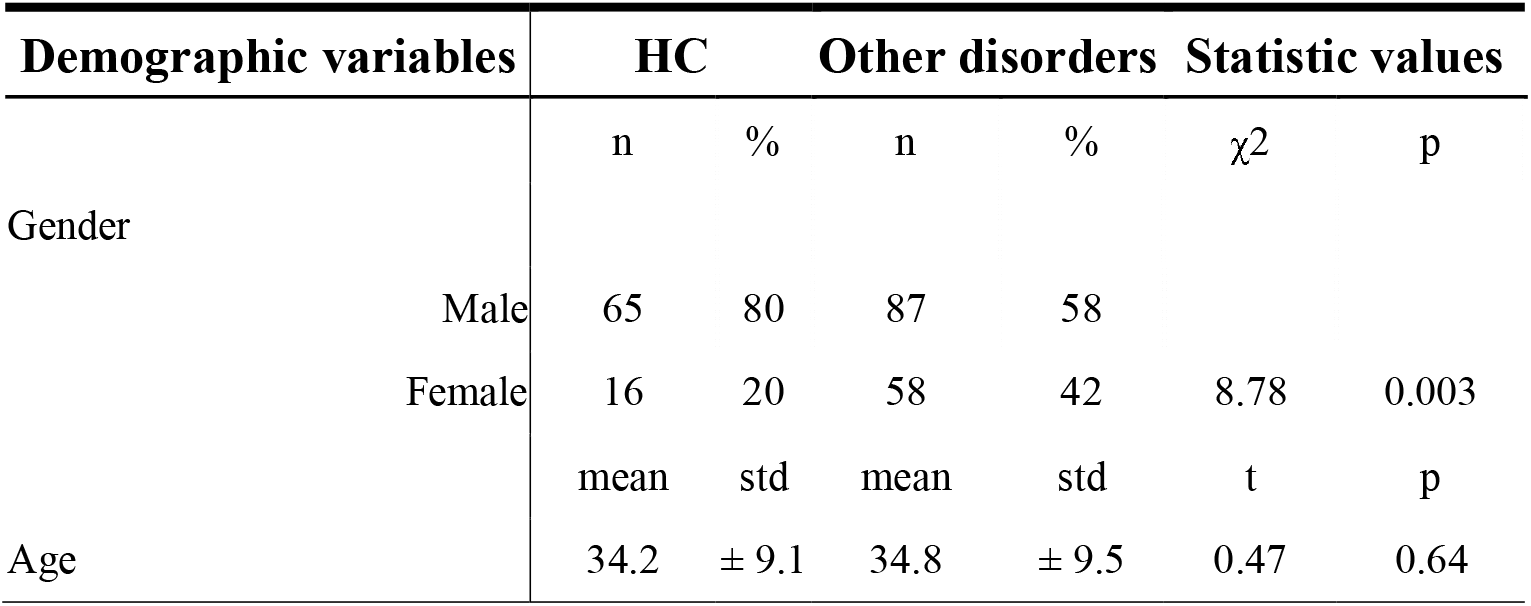
Demographic information of UCLA-CNP datasets.

**Table S8.** Demographic information of the replication cohort.

| Demographic variables |  | HC |  | CUD |  | Statistic values |  |
| --- | --- | --- | --- | --- | --- | --- | --- |
| | | n | % | n | % | $\chi^2$ | p |
| Gender | Male | 65 | 80 | 71 | 87 | 0.77 | 0.38 |
|  | Female | 16 | 20 | 11 | 13 |  |  |
|  |  | mean | std | mean | std | t | p |
| Age | | 34.2 | $\pm 9.1$ | 35.7 | $\pm 8.2$ | -1.12 | 0.26 |

